# Pre-HCT Resistome Disruption Predicts ESBL Gene Expansion in Pediatric Transplant Recipients: A Prospective Multi-Center Study

**DOI:** 10.64898/2026.01.20.26344466

**Authors:** Marygrace Duggar, Yilun Sun, Davide Leardini, Qidong Jia, Edoardo Muratore, Ronald H. Dallas, Jose Ferrolino, Anju Cherian, Simone Cesaro, Maura Faraci, Jowita Fraczkiewicz, Marek Ussowicz, Janet A. Englund, Hana Hakim, Randall T. Hayden, Eileen J. Klein, Joshua Wolf, Gabriela Maron, Li Tang, Riccardo Masetti, Elisa B. Margolis

## Abstract

**Background:** Infections are the leading cause of non-relapse mortality in pediatric hematopoietic cell transplant (HCT) recipients. Up to 90% of bacteremias in these patients originate from gut microbiome organisms. However, selection for resistance genes, such as Extended-spectrum β-lactamase (ESBL), in these patient’s gut microbiomes remains poorly understood.

**Methods:** Stools were prospectively collected from pediatric HCT recipients at multiple centers (n=133 patients, five centers) on the day of HCT, the day of neutrophil engraftment, and 30 days post-HCT. Bacterial DNA was isolated and sent for shotgun metagenomic sequencing. Antibiotic resistance genes were identified using the MEGARes database. Associations between ESBL gene abundance changes and antibiotic exposure were examined using univariate and Inverse Probability of Treatment Weighting linear regression models with covariate balancing propensity scores.

**Results:** Pre-existing gut resistome disruption at the time of HCT showed a stronger correlation with ESBL gene expansion than post-transplant antibiotic exposure. Specifically, patients with greater baseline resistome distance from healthy children showed increased ESBL genes during the neutropenic period. Post-transplant β-lactam exposure (total or ESBL-cleavable) did not correlate with increases in ESBL genes in already-colonized patients. However, aminoglycosides and anaerobic active antibiotics were associated with acquisition of new ESBL organisms during the neutropenic period, while pre-existing microbiome disruption primarily drove selection of resistant bacteria already present.

**Conclusions:** These findings indicate that antibiotic stewardship before HCT, in addition to reducing the use of anaerobic active antibiotics during early transplant, may be necessary to prevent ESBL-related infections in pediatric transplant recipients.

**Lay Summary:** Infections are the leading cause of death after HCT, and recently the role of the gut microbiome in harboring dangerous bacteria has been highlighted. This study aims to understand multidrug resistant bacteria changes in the gut microbiome early after HCT.

## Introduction

Bacterial infections are the leading cause of non-relapse mortality in hematopoietic stem cell transplant (HCT) recipients^1^, with extended-spectrum β-lactamase (ESBL)-producing bacteria^2^ representing a particularly lethal threat with mortality rates of 15 to 45%^3^ in immunocompromised patients. A major source of these infections is the patient’s gut microbiome, which serves as a detectable reservoir for up to 90% of bacteremia cases in pediatric HCT recipients^4^. Pre-transplant conditioning both damages the gut epithelium to allow for increased bacterial translocation^5,6^ and disrupts the protective microbial communities that normally confine resistant organisms to minor constituents of the intestinal community^7,8^. The neutropenic period following transplant requires prolonged broad-spectrum antibiotics, which would be expected to exert selective pressure favoring ESBL genes in the gut microbiome.

Despite the clear association between broad-spectrum antibiotic exposure and resistance emergence in immunocompetent patients^9^, the specifics of which antibiotics (e.g. specific antibiotics, which treatment courses, impact of short empiric courses or prophylaxis) select for ESBL genes in the gut microbiomes of pediatric HCT recipients remain poorly understood. Understanding these dynamics is critical for developing antibiotic stewardship and targeted interventions to prevent ESBL colonization and subsequent life-threatening infections in these vulnerable population.

Antibiotics have strong bystander effects, acting on bacteria in the gut microbiome that are not the target of the treatment^10^. Resistance gene expansion in the gut can occur through two primary mechanisms: selection and enrichment of pre-existing resistant bacteria already present in the microbiome, or acquisition and colonization by new resistant organisms from external sources. Exposure to these antibiotics can drive both mechanisms in multiple ways. The most straightforward is direct selection, where bacteria exposed to a particular antibiotic agent become resistant to that antibiotic^11^. Some antibiotic resistance genes coexist on plasmids or can be coregulated, and so exposure to one antibiotic can indirectly select for resistance to another class of antibiotics^11^. Additionally, radiation and chemotherapy, which are part of transplant conditioning regimens, put stress on the bacteria, altering their physiology and inducing DNA mutations that could result in increased antibiotic resistance or lead to the loss of keystone members of the gut microbial community^12^. Many of these antibiotic resistance genes impose a metabolic cost, allowing susceptible bacteria to dominate in an intact microbiome through competitive exclusion^13^. However, antibiotic-mediated microbiome disruption can open ecological niches within the community, leaving the community vulnerable to resistant organism colonization and expansion.

With all the different concomitant exposures that could alter the selection, maintenance and fitness of resistance genes during early transplant, it is unknown whether specific antibiotic exposures drive ESBL gene expansion in pediatric HCT recipients. We performed a retrospective cohort study of prospectively collected stool samples to characterize the resistome-the complete collection of antibiotic resistance genes-in two cohorts from the United States and Europe during early transplant to capture the influence of antibiotics in diverse clinical practices and populations.

## Methods

### Patient population

Samples were prospectively collected from two cohorts of pediatric HCT recipients. From November 2015 to January 2020 94 patients underwent allogeneic HCT at St. Jude Children’s Research Hospital (Memphis, TN, USA). From June 2022 to November 2023 39 patients underwent allogeneic HCT at 4 centers in Europe (IRCCS Azienda Ospedaliero-Universitaria di Bologna (European organizing center), Azienda Ospedaliera Universitaria Integrata of Verona, Wrocław Medical University, and IRCSS Istituto Giannina Gaslini). All participants provided stool samples before HCT, 30 days after HCT, and/or after neutrophil engraftment (NE). The study was approved by Institutional Review Boards at participating centers with informed consent (and when applicable assent) obtained. Participant characteristics can be found in table 1. Briefly, the median age for all participants was 8.2 years (range 0.00-29.4). Indications for transplantation included acute myeloid leukemia (43.6%), acute lymphoblastic leukemia (ALL) (35.3%), lymphoma (3.8%), myelodysplastic/myeloproliferative syndrome (4.5%), nonmalignant disease (12.0%). Most patients received either myeloablative (47.4%) or reduced intensity (51.9%) conditioning. For comparison, we used age-matched de-identified leftover research stool samples from 347 healthy children and 91 children undergoing ALL treatment. These samples were obtained through research protocols previously approved by Seattle Children’s Hospital and St Jude’s IRB.

**Table 1.** Patient Characteristics.

| Characteristic | European (N = 39) | United States (N = 94) | P-value |
| --- | --- | --- | --- |
| Years collected | 2015-2019 | 2022-2023 |  |
| Age at Transplant | 8.1 [0.0, 29.4] | 8.4 [0.8, 18.3] | 0.865 |
| Gender |  |  | 0.908 |
| Female | 17 (43.6%) | 42 (44.7%) |  |
| Male | 22 (56.4%) | 52 (55.3%) |  |
| Race |  |  | 0.145 |
| Asian | 0 (0.0%) | 2 (2.1%) |  |
| Black | 0 (0.0%) | 7 (7.4%) |  |
| Multiple | 0 (0.0%) | 5 (5.3%) |  |
| Other | 0 (0.0%) | 2 (2.1%) |  |
| White | 39 (100.0%) | 78 (83.0%) |  |
| Ethnicity |  |  | <0.001 |
| Hispanic | 1(2.6%) | 36 (38.3) |  |
| Non-hispanic | 38 (97.4%) | 58 (61.7%) |  |
| Diagnosis |  |  | <0.001 |
| Acute myeloid leukemia | 7 (17.9%) | 51 (54.3%) |  |
| Acute lymphoblastic leukemia | 12 (30.8%) | 33 (35.1%) |  |
| Lymphoma | 1 (2.6%) | 3 (3.2%) |  |
| Myelodysplastic syndrome | 3 (7.7%) | 3 (3.2%) |  |
| Other malignancy<sup>1</sup> | 2 (5.1%) | 4 (4.3%) |  |
| Nonmalignant disease<sup>2</sup> | 14 (35.9%) | 0 (0.0%) |  |
| Prior Transplant |  |  | <0.001 |
| No | 23 (59.0%) | 85 (90.4%) |  |
| Yes | 16 (41.0%) | 9 (9.6%) |  |
| Donor Match Status |  |  | <0.001 |
| Haploidentical | 7 (17.9%) | 54 (57.4%) |  |
| Matched related | 5 (12.8%) | 14 (14.9%) |  |
| Matched unrelated | 27 (69.2%) | 25 (26.6%) |  |
| Mismatched unrelated | 0 (0.0%) | 1 (1.1%) |  |
| Conditioning Intensity |  |  | <0.001 |
| Myeloablative | 31 (79.5%) | 32 (34.0%) |  |
| Non-Myeloablative | 0 (0.0%) | 1 (1.1%) |  |
| Reduced Intensity | 8 (20.5%) | 61 (64.9%) |  |
| Irradiation |  |  | 0.003 |
| None | 28 (71.8%) | 45 (47.9%) |  |
| Total Body Irradiation | 10 (25.6%) | 22 (23.4%) |  |
| Total Lymphoid Irradiation | 1 (2.6%) | 27 (28.7%) |  |
| Graft Source |  |  | 0.417 |
| Bone Marrow | 15 (38.5%) | 37 (39.4%) |  |
| Cord Blood | 1 (2.6%) | 0 (0.0%) |  |
| Peripheral Blood Mononuclear Cells | 23 (59.0%) | 57 (60.6%) |  |
| T Cell Depletion |  |  | 0.006 |
| No | 11 (28.2%) | 51 (54.3%) |  |
| Yes | 28 (71.8%) | 43 (45.7%) |  |
| Type of T Cell Depletion |  |  | <0.001 |
| Anti-thymocyte Globulin | 28 (71.8%) | 0 (0.0%) |  |
| CD3 Depletion | 0 (0.0%) | 1 (1.1%) |  |
| CD34 Enrichment | 0 (0.0%) | 1 (1.1%) |  |
| CD45RA Depletion | 0 (0.0%) | 3 (3.2%) |  |
| CD45RA Depletion/CD34 Enrichment | 0 (0.0%) | 30 (31.9%) |  |
| CD45RA Depletion/TCR AB Depletion | 0 (0.0%) | 7 (7.4%) |  |
| None | 11 (28.2%) | 51 (54.3%) |  |
| TCR AB Depletion | 0 (0.0%) | 1 (1.1%) |  |
| Gut GVHD |  |  | 0.338 |
| 0-1 | 32 (82.1%) | 83 (88.3%) |  |
| 2-4 | 7 (17.9%) | 11 (11.7%) |  |

<sup>1</sup>Other malignancy contains acute biphenotypic leukemia, T cell large granular lymphocytic leukemia, renal cell carcinoma, and neuroblastoma.
<sup>2</sup>Nonmalignant diseases include Bone Marrow Aplasia, Wiscott-Aldrich syndrome, Hurler syndrome, Severe Combined Immunodeficiency, Juvenile Idiopathic Arthritis, CTLA4 deficiency, Severe Aplastic Anemia, Leukocyte Adhesion Deficiency type 1, congenital dyskeratosis, Fanconi anemia, and chronic granulomatous disease.

Demographic data, transplant details, and antibiotic exposure were collected from electronic medical records. Antibiotic exposure was calculated as cumulative daily doses, counting each antibiotic (including prophylaxis) administered on a given day separately, even if from the same class. Antibiotics were classified by spectrum (e.g. anaerobic, ESBL-susceptible β-lactams) according to Antibiotic and β-lactam Spectrum Index^14^.

### Resistome analysis

Stool samples were frozen at −80°C. DNA was extracted using the QIAamp DNA Stool Mini Kit (Qiagen) and sequenced on Illumina NovaSeq (100-bp paired-end reads, ~25 million reads per sample (supplemental figure 1)). Antibiotic resistance genes (ARGs) were identified using the AMR++ v3 pipeline^15^ with MEGARes V3.0 database, filtered to included only genes with >80% gene length coverage. ARGs were classified by mechanism and antibiotic class. Supplemental table 5 lists specific genes identified as ESBL genes. ARG abundance was normalized as natural log of reads per kilobase (gene length) per million reads (lnRPKM+1).

### Statistical analysis

We measured the presence and relative abundance of ESBL genes at three time points: prior to HCT, 30 days after HCT, and within 5 days after NE (defined as the first of 3 consecutive days where absolute neutrophil count was >500mm^3^). Within-participant changes were evaluated using Wilcoxon signed-rank tests (abundance) or McNemar’s test (prevalence). P-values were false discovery rate (FDR) adjusted.

Associations between ESBL gene abundance changes and antibiotic exposure were examined using univariate and Inverse Probability of Treatment Weighting linear regression models with covariate balancing propensity scores^16^ based on age, sex, underlying diagnosis (leukemia v non-leukemic), graft source, and GVHD status. Gene presence changes were categorized as “Newly Present”, “Lost Detection”, “Remained Present”, or “Remained Absent”, with antibiotic exposure comparisons using permutation tests^17^. Additionally, linear regression was used to evaluate the association between changes in Bray-Curtis dissimilarity to healthy cohort centroids and changes in ESBL gene abundance. All analyses were performed in R.

## Results

### ESBL Gene Dynamics Across Early Transplant

Across two cohorts of pediatric patients undergoing HCT at six transplant centers, there were differences in the proportion of participants with detectable ESBL genes over the first 30 days after transplantation (Fig 1a). The United States cohort had higher baseline ESBL colonization prevalence than the European cohort which declined post-transplant, whereas the European cohort showed increasing prevalence through day 30.

**Figure 1:**
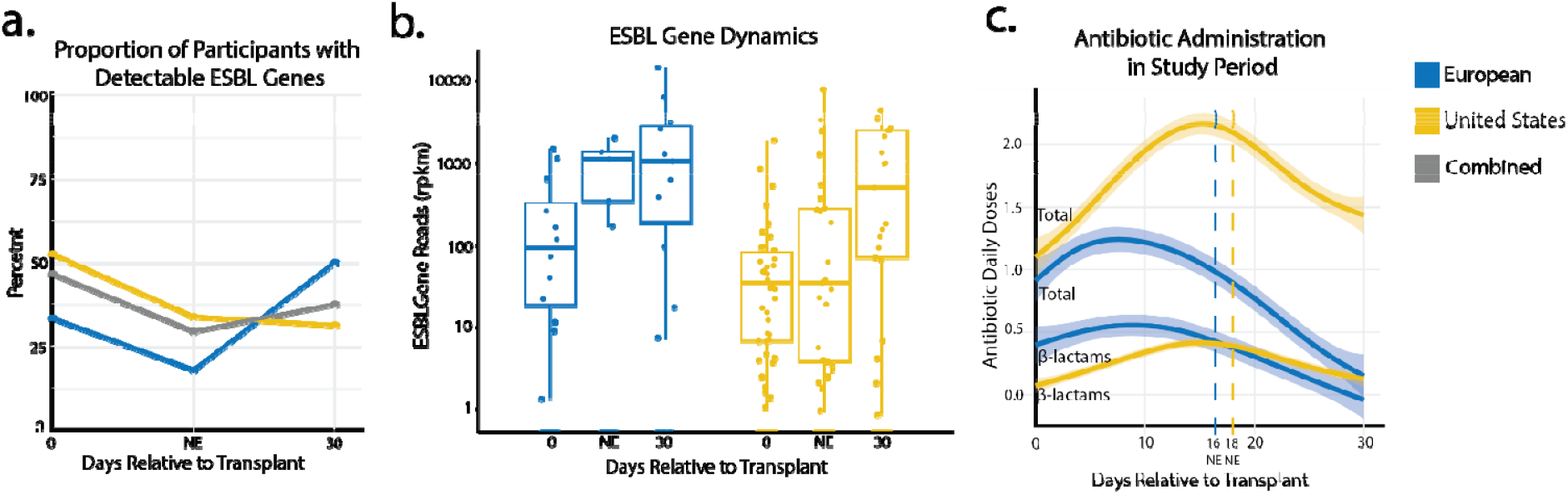
Cohort-specific ESBL gene and antibiotic exposure dynamics during early transplant. A) Prevalence of ESBL genes at each time points. The United States cohort had higher baseline prevalence that decreased over time, while the European cohort showed increasing prevalence. B) ESBL genes abundance normalized to sequencing epth (rpkm-reads per kilobase of gene length per million sequencing reads) increased between different intervals in the cohorts. C) Daily antibiotic doses differed between cohorts. The United States cohort received more total daily doses of antibiotics due to prophylactic regimens, whereas the European cohort received more β-lactams during the neutropenic window. Neutrophil engraftment (NE) timing was similar between cohorts.

Among participants with detectable ESBL genes, gene burden increased for both cohorts early after transplant (Fig 1b), reflecting either expansion of pre-existing ESBL-producing bacteria or acquisition of new ESBL-harboring organisms. However, the kinetics of increases differed between cohorts. The European cohort showed an increase between transplant and NE which was maintained at day 30. In contrast, the United States cohort showed no increase in genes during the neutropenic period but demonstrated selection for ESBL genes between NE and day 30.

### Antibiotic Exposure and Selection for ESBL Genes

Antibiotic exposure patterns differed substantially between cohorts. The United States cohort received significantly more total antibiotic days compared to the European cohort (Fig 1c), likely due to difference in prophylactic regimens. St Jude’s standard of care included continuous antibiotic prophylaxis with trimethoprim-sulfamethoxazole for *Pneumocystis jiroveci* Pneumonia prevention throughout the transplant period. European centers did not use prophylactic antibiotics, administering only empiric and therapeutic antibacterial courses. The European cohort received significantly more daily doses of β-lactams compared to the United States cohort, primarily before NE. The ESBL gene burden temporal trends appear to track with this differential timing of β-lactam exposure in each cohort shown in fig 1b. Full antibiotic exposure details can be found in supplemental table 1.

Despite these apparent trends, univariate logistic regression exposure showed no correlation between β-lactams, ESBL-susceptible β-lactams, antibiotics with anaerobic activity or total antibiotic daily doses and an increase in ESBL genes for either cohort early after transplant. Prior to engraftment (day 0 to NE) only, aminoglycoside exposure was significantly associated with increased ESBL gene expansion in the European cohort (fig 2a, supplemental table 2). Aminoglycosides were administered too infrequently in the United States cohort for this comparison to be performed. Comparing antibiotic exposures in the first 30 days after HCT yielded no significant associations for either cohort (fig 2b, supplemental table 3).

**Figure 2:**
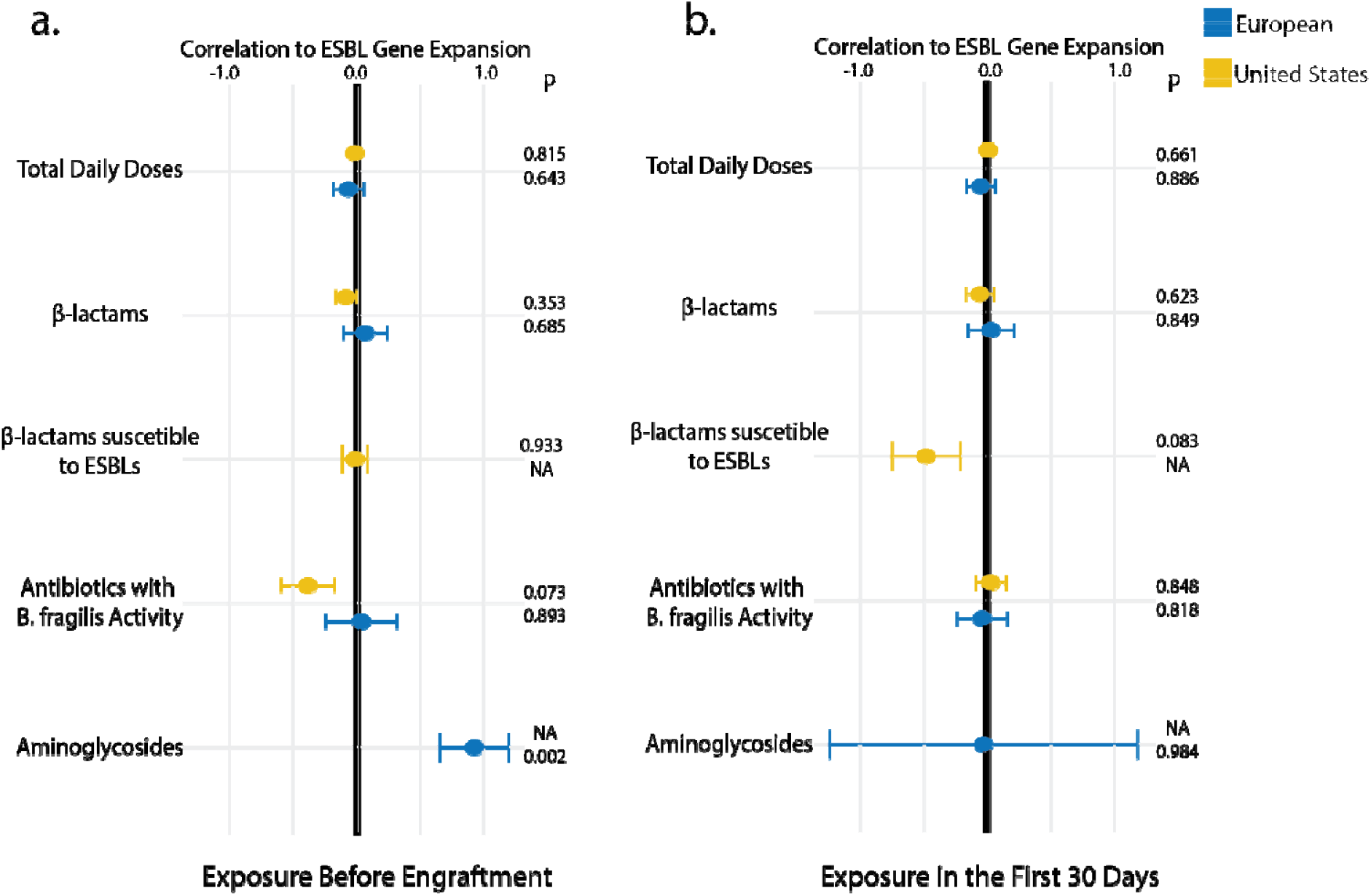
Associations of antibiotic expsoures and ESBL gene abundance changes. A) Before engraftment only aminoglycoside exposure was associated with increases in ESBL genes from transplant. Aminoglycosides were administered too infrequently in the United States cohort for their impact to be analyzed. Surprisingly, neither cohort had correlations with total antibiotic doses or daily doses of beta-lactams, or beta-lactams cleavable by ESBL enzymes, or all antibiotics with anaerobic activity and changes in ESBL abundance. B) 30 days after transplant none of the antibiotic exposures correlated with changes in ESBL abundance for either cohort. P-values adjusted for multiple comparisons and confounding variables using CBPS weighting.

To distinguish how antibiotics alter acquisition of new resistant organisms, we compared the participants who remained negative for ESBL genes to those who converted from negative to detectable for each cohort (fig 3). Participants converting from ESBL-negative to ESBL-positive likely represent new acquisition events, which may be masked in the previous analyses by increases in ESBL gene abundance among already-colonized patients. Prior to engraftment (day 0 to NE), European cohort participants who acquired microbes with ESBL genes received significantly more days of aminoglycosides (fig 3e) and antibiotics with activity against *B. fragilis* (a marker of anaerobic activity) (fig 3d) than patients who remained negative. In the United States cohort, acquiring ESBL genes was associated with daily doses of β-lactams (fig 3b) and antibiotics with anaerobic activity (fig 3d). No significant associations were observed between acquired microbes with ESBL and antibiotic exposure when comparing between day 0 and day 30 (supplemental figure 2).

**Figure 3:**
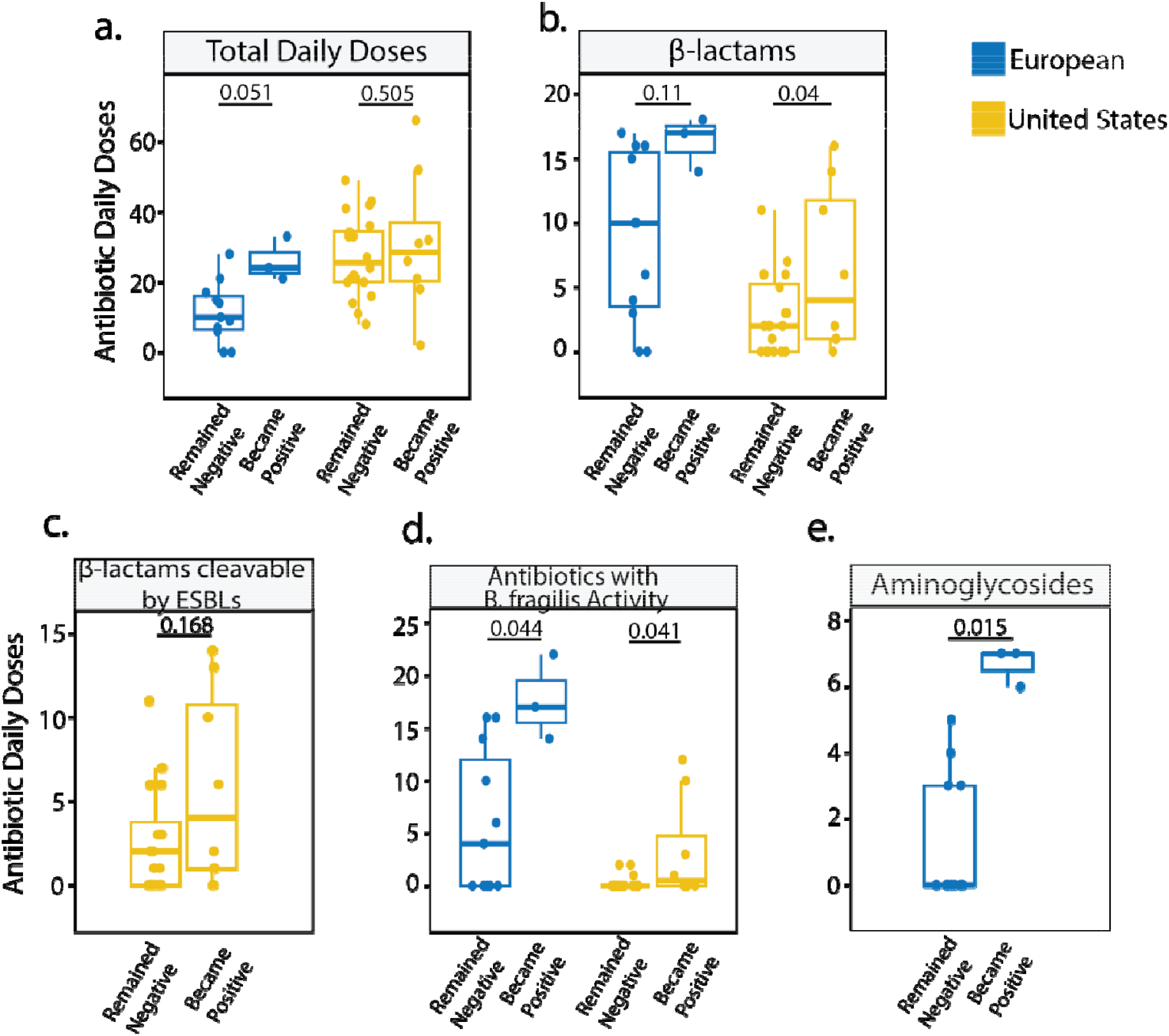
Antibiotic exposures associated with new ESBL gene acquisition. Among participants who were ESBL-negative prior to HCT, antibiotic exposures during neutropenia were compared for those who remained negative versus those who acquired ESBL genes. A) Total antibiotic doses did not differ significantly between groups in either cohort. B) Total β-lactam exposure was higher in new ESBL acquisition in the United States cohort but not the European cohort. C) ESBL-cleavable β-lactams were not higher in the new ESBL acquisition United States cohort; insufficient exposure in the European cohort precluded analysis. D) Antibiotics with B. fragilis coverage (a proxy for anaerobic activity of an antibiotic) was increased for new ESBL acquisition in both cohorts. E) Aminoglycoside exposure was higher in the new ESBL acquisition European cohort; insufficient exposure in the United States cohort precluded analysis. P values were calculated using permutation tests and adjusted using the Benjamini–Hochberg false discovery rate method.

### Pre-existing Resistome Disturbance Correlates with ESBL Gene Dynamics

The inability of antibiotic exposure alone to explain changes in ESBL gene presence and expansion led us to consider broader patterns of microbial disruptions in HCT patient gut microbiomes. We compared the composition of antibiotic resistance genes (resistome) of the United States and European cohorts with those from age-matched healthy children and patients undergoing first-line chemotherapy for acute lymphoblastic leukemia (ALL). Both HCT cohorts showed significant resistome differences from healthy children, even in the samples prior to HCT (Fig 4a). Children early in ALL chemotherapy demonstrated resistome compositions between healthy and HCT cohorts but remained closer to the healthy children than the HCT recipients. In comparison, very little shift was seen in the resistomes during early transplantation.

**Figure 4:**
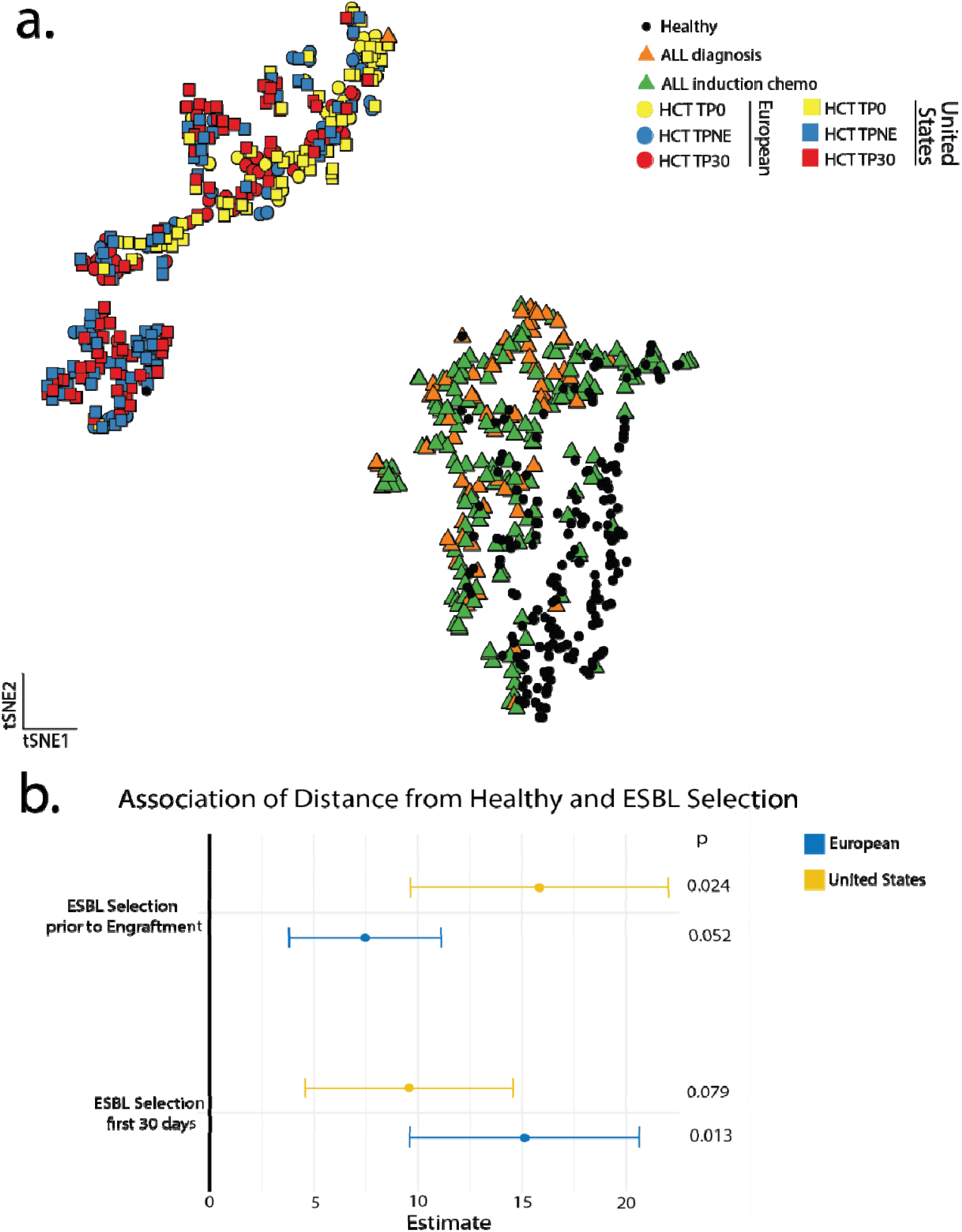
Preexisting disruption in HCT recipients compared to healthy children and leukemia patients. A) Resistome composition of HCT recipients are displayed according to their similarity (Bray-Curtis Distance), with samples with more similar resistance genes closer together on the tSNE plot. The three timepoints (day 0, neutrophil engraftment (NE), and day 30) of both HCT cohorts cluster together, indicating minimal resistome changes during early transplant despite intensive antibiotic exposure. In contrast, HCT resistomes are highly distinct from both healthy age-matched children and children with acute lymphoblastic leukemia (ALL) sampled early in chemotherapy. B) Bray-Curtis distance from the age-matched healthy-cohort centroid of the resistome at baseline is associated with increased ESBL gene abundance at subsequent timepoints (neutrophil engraftment and day 30). Associations were assessed using univariate logistic regression with propensity score weighting.

We compared whether the degree of disturbance (Bray Curtis dissimilarity of sample prior to transplant to the centroid of the healthy cohort, illustrated in supplemental figure 3) was associated with increased ESBL gene abundance in both cohorts (fig 4b, supplemental table 4). According to univariate linear regression, the degree of disturbance in sample prior to transplant correlated with selection of ESBL genes during early transplant (significant at NE in the United States cohort and at post-transplant day 30 in European cohort), indicating that baseline disturbance is an important predictor of increases in ESBL-harboring bacteria.

## Discussion

Pediatric transplant recipients show selection for ESBL genes early after transplant, but the timing and kinetics differ substantially between cohorts. These differences reflect variation in selective pressures, including antibiotic exposure, prophylaxis protocols, and conditioning/graft features. This early post-transplant period presents the greatest risk for life-threatening infections from highly resistant bacteria. By examining the gut reservoir, we were able to identify a broader microbial context which influences selection for these organisms before they cause infection. Pre-existing microbiome disruption appeared to correlate with ESBL gene expansion more strongly than post-transplant antibiotic exposure. We anticipated that β-lactam exposure would directly select for ESBL genes during early transplant, but instead the degree of resistome dissimilarity from healthy children prior to transplant was more significantly associated with ESBL gene increases. Total β-lactam or anaerobic antibiotic exposure did not correlate with increases, but the latter did correlate with new acquisition of ESBL genes. These observations raise the possibility that antibiotics during early transplant can lower the colonization barrier for ESBL-harboring bacteria, but that pre-existing resistome disruption before HCT has a larger impact on ESBL selection. These findings highlight that antibiotic stewardship interventions before HCT warrant further investigation as a potential strategy for infection prevention in pediatric transplant recipients.

Timing of ESBL gene burden differed between cohorts. The European cohort showed increases between day 0 and NE that persisted through day 30. The United States cohort showed increases only between engraftment and day 30. These patterns reflect center-specific variables that shape resistance gene dynamics. Conditioning regimens^18^, antibiotic prophylaxis protocols^19^, transplanting protocols^20^, and indications for transplant^21^ can shape gut microbial communities and vary substantially across centers. In pediatric patients, this variability is further complicated by the extensive developmental changes of the gut microbiome throughout childhood^22,23^. These cohort-specific dynamics underscore the challenge of generalizing findings from single center studies^24^.

B-lactam exposure did not correlate with ESBL gene expansion overall, suggesting that direct selection alone may not fully explain resistance dynamics. Neither total β-lactams nor ESBL-susceptible β-lactams correlated with ESBL gene increases during the neutropenic window, pointing to alternative mechanisms. This could reflect several non-mutually exclusive possibilities: a) indirect selection through community disruption, b) short-lived direct selection could not be captured with our sampling frequency, or c) pre-existing resistance saturation masking selection signals. In the European cohort, aminoglycoside exposure correlated with ESBL acquisition and selection, possibly through linkage disequilibrium, where aminoglycoside modifying enzymes co-occur with ESBL on the same plasmids (or organisms)^25,26^. However, aminoglycosides resistance genes did not have similar kinetics to ESBL genes (supplemental figures 4 and 5). This could reflect community-level effects, where aminoglycosides disrupt specific gut microbiome members and create niches for ESBL-harboring organisms, though partial co-selection of only a subset of aminoglycoside genes on mobile genetic elements carrying ESBLs also remains plausible.

While β-lactam exposure was associated with new ESBL gene detection in the United States cohort, it did not correlate with increases in ESBL gene abundance among already-colonized patients in either cohort. This distinction between acquisition (colonization by new organisms) and selection (expansion of existing organisms) is consistent with a model where β-lactams may lower the colonization barrier for ESBL-producing bacteria without necessarily providing strong selective pressure for their subsequent expansion during the neutropenic period. This pattern extends to other antibiotic classes that contribute to indirect antibiotic resistance gene selection through community-level effects, by disrupting gut microbiome members and allowing new acquisition of multi-drug-resistant organisms (MDROs). Both cohorts showed this pattern: participants converting from ESBL absent to present received significantly more anaerobic antibiotics than those who maintained ESBL absent status (Fig 1d). While alternative explanations remain possible, these observations suggests that anaerobic antibiotic stewardship may reduce new MDRO acquisition by preserving colonization resistance in high-risk patients.

Both HCT cohorts showed markedly distinct resistome compositions (compared to healthy children) even in samples collected prior to transplant, suggesting that much of the resistome disturbance may have occurred before HCT rather than during the intensive antibiotic exposure period following transplant. Relatively modest shifts occurred in the resistomes during early transplantation despite heavy antibiotic usage. This pattern could reflect some combination of: a) saturation of resistance genes prior to HCT, b) insufficient sampling frequency to capture rapid and short-lived dynamics, or c) resilience of resistant populations once established. None of the children in our study were likely antibiotic naïve before transplant, as children undergoing transplant for malignancy likely experienced months of previous antibiotic, chemotherapeutic, and other medical exposures that shaped their gut microbiomes. Children early in ALL treatment demonstrated resistome compositions distinct from both healthy and HCT cohorts but did not completely explain the resistome disturbance in the HCT cohorts likely because HCT cohorts were enriched for patients with high-risk disease (e.g. AML) while most ALL patients have favorable outcomes not requiring HCT.

Pre-existing microbiome disruption at the time of HCT strongly predicted ESBL gene expansion during early transplant. This pattern is compatible with several interpretations: earlier antibiotic exposures exert disproportionate impact on the gut resistome or alternatively, cumulative exposures creating resilient resistance reservoirs. We speculate that gut microbiomes may reach a saturation of antibiotic resistance genes (due to fitness costs), and this saturation may have been reached prior to HCT, though this hypothesis requires direct testing. If true, concentrating antibiotic stewardship on earlier (prior to transplant) courses could reduce the antibiotic-resistant organisms that transplant recipients’ harbor.

While our findings suggest that pre-HCT resistome status may be more predictive of ESBL gene dynamics than post-transplant exposure, these observations require validation in prospective interventional studies. Importantly, our study design cannot definitively establish casual relationships between pre-HCT resistome status and subsequent expansion of ESBL genes. Unmeasured confounders, including illness severity, duration of cancer therapy, and genetic susceptibility to microbiome disruption, may contribute to the observed associations. While suggestive, the correlative nature of these observations necessitates cautious interpretation. The clinical impact of modifying pre-HCT microbiomes (and their resistomes) on actual infection outcomes remains unknown. Future studies linking resistome changes to ESBL bloodstream infections and evaluating targeted pre-HCT interventions are needed to translate these findings into evidence-based practice.

Resistome studies provide a more complete picture than epidemiology studies of patients that go onto have infections with multi-drug-resistant organisms but still have a number of limitations and caveats. Identification of ARGs was limited by the availability of documented sequences. MEGARes 3.0 is a comprehensive database of reported ARG sequences, but it’s possible that unreported sequences were present and not detected. Also, the coverage of small point mutations of existing genes to avoid antibiotics may not be well detected. Furthermore, metagenomic detection does not confirm phenotypic resistance or bacterial viability. We cannot determine whether the ESBL genes were actively expressed or conferred elevated minimum inhibitory concentrations to β-lactam antibiotics. This is the trade-off for metagenomic approaches that capture resistance potential from both culturable and difficult to grow organisms and our longitudinal within-patient comparisons help control for individual variation in the relationship between genotypic and phenotypic resistance. Previous studies in HCT recipients have validated that metagenomic detection of organisms in the gut microbiota strongly predicts subsequent clinical bloodstream infections^27–30^. Nevertheless, future work directly linking resistome changes to infection outcomes and antimicrobial susceptibility testing would strengthen the translational implications of this approach.

We specifically designed this study to be able to capture the heterogeneity across centers (in clinical practices, local resistance epidemiology, conditioning and prophylaxis protocols, transplant indications) but the study was not powered to be able to interpret how these components interact with the selective pressure of antibiotics. Although propensity score weighting allowed us to balance measured covariates between groups, residual confounding from unmeasured center-level factors (conditioning protocols, prophylaxis practices, resistance epidemiology and temporal trends) remains a limitation of this observational study. Finally, we choose timepoints (baseline, NE, day 30) based on when clinical interventions directed at the gut microbiome may be considered and when patients receive the highest burden of antibiotics post-transplant, but the cohort-specific patterns suggest that generalizability may require more in depth sampling frequency.

There is great clinical interest in gut microbiome modulation to prevent HCT complications^31,32^, however interventions with live bacteria during neutropenia are strongly discouraged due to the risk of severe infection. Therefore, in previous work, focus has been on intervening after NE^33,34^. Our findings on pre-transplant resistome disruption, together with previous observations linking pre-transplant diversity to post-transplant outcomes in children^35,36^, are consistent with a potential role for pre-HCT microbiome interventions (such as FMT or other targeted therapies) though substantial uncertainty around safety and efficacy remain. Additionally, the association between antibiotics with anaerobic activity and ESBL-harboring bacteria acquisition supports continued emphasis on antibiotic stewardship efforts both before and after HCT. Antibiotics with anaerobic activity have the ability to target healthy bacteria within the gut microbiome and open a potential niche for resistant bacteria to colonize^37^. Our data suggests these disruptions can be long-lasting, and antibiotic exposure throughout cancer care has impacts on ESBL gene expansion early in transplant (fig 3, supplemental tables 2 and 3). Antibiotics remain life-saving therapy, and our findings reinforce the importance of choosing narrow spectrum agents whenever clinically appropriate throughout cancer therapy and HCT^38^. Antibiotic stewardship before HCT, in addition to reducing the use of anaerobically active antibiotics during early transplant, may prevent ESBL-related infections in pediatric transplant recipients.

## Data Availability

Data not included in the manuscript will be available by request to the corresponding authors. All metagenomic data will be uploaded to the Sequence Read Archive accession number PRJNA1391538.

