## supplemental methods, figures, and tables for "Pre-HCT Resistome Disruption Predicts ESBL Gene Expansion in Pediatric Transplant Recipients: A Prospective Multi-Center Study"

**Patient population**

The de-identified leftover research stool samples from 347 healthy children were collected at Seattle Children’s Hospital between 2011-2016 with a single sample per child and designated as non-human samples by the St Jude Children’s Research Hospital IRB.

The samples from children with ALL were collected at St. Jude Children’s Research Hospital between 2012-2015. Samples from these children were collected at the time of ALL diagnosis and then longitudinally through induction and consolidation chemotherapy. The collection and patient characteristics were described previously^1^.

**Resistome analysis**

Antibiotic resistance genes were identified using MEGARes v3.0. Briefly, sequence reads were trimmed and filtered using Trimmomatic V0.39^2^. Quality trimmed reads were aligned to Human GRCh38 genome using BWA-MEM V0.7.17-r1188, and human reads were removed using Samtools V1.17^3^.

**Supplemental Figures:**

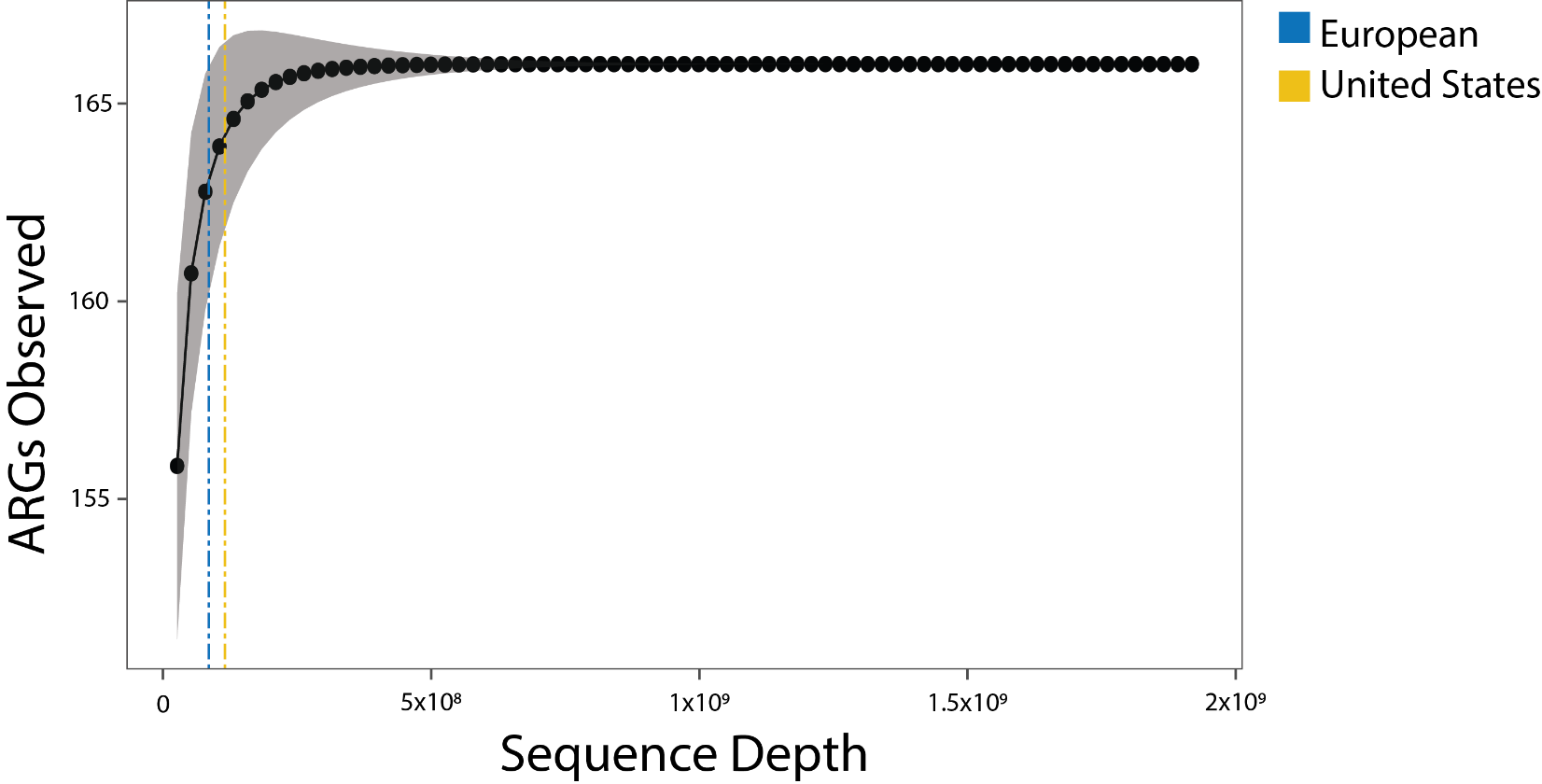

**Supplemental figure 1: ARG accumulation curves.** Number of antibiotic resistance genes (ARGs) observed in the data is related to the total sequencing depth. Dotted lines represent median sequencing depth for each cohort and indicate that sequencing depth was sufficient to capture >98% of the ARGs present in the samples.

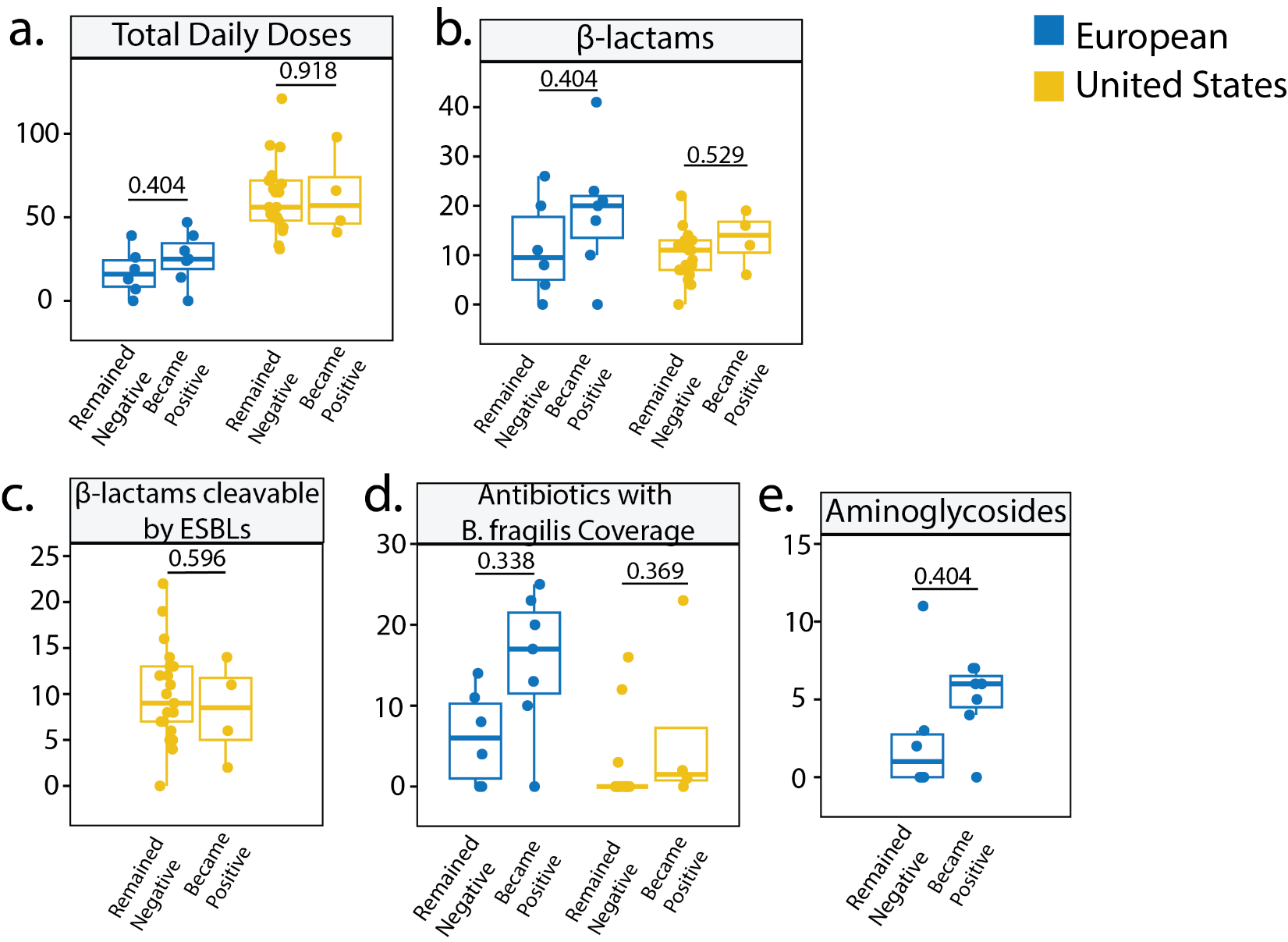

**Supplemental Figure 2: day 0 to day 30 daily doses of antibiotics between participants who acquired ESBL genes versus those who remained negative.** Among participants who were ESBL-negative prior to HCT, antibiotic exposures in the first 30 days post HCT were compared for those who remained negative versus those who acquired ESBL genes. a-e.) None of the assessed antibiotics dosing differed in either cohort. Analysis of ESBL cleavable beta-lactams in the European cohort and aminoglycosides in the United States cohort were not done due to very rare administration of those agents. P values were calculated using permutation tests and adjusted using the Benjamini–Hochberg false discovery rate method.

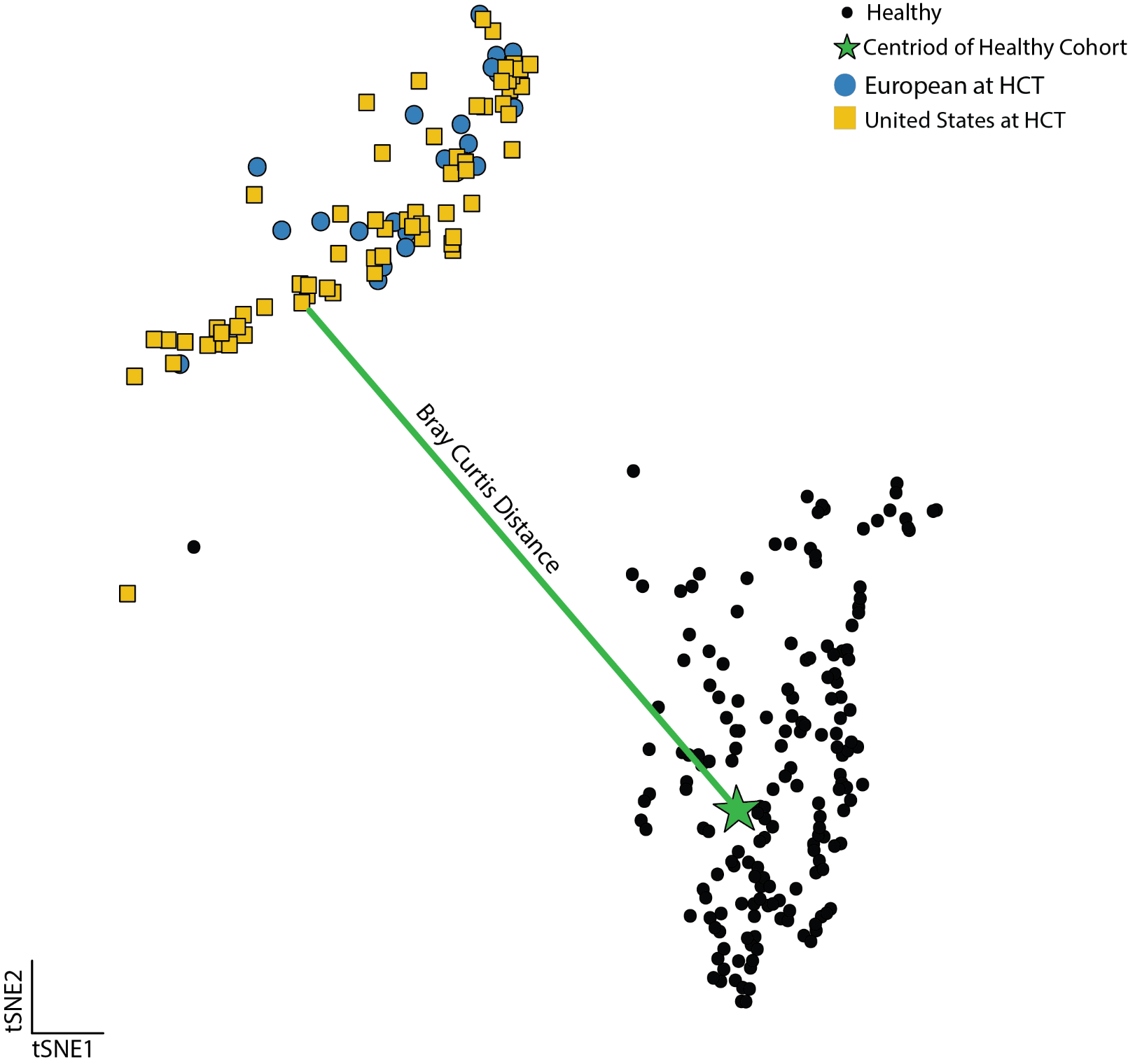

**Supplemental Figure 3: Bray Curtis distance from Healthy Centroid.** To determine the effect of global resistome changes on the likelihood of accumulating new ESBL genes, we calculated the Bray-Curtis distance of the baseline sample from each participant to the centroid of the healthy cohort. The larger the bray curtis distance the more distinct the sample is from the centroid value. The figure above depicts a visual of the bray curtis distance between the centroid and one participant in United States cohort.

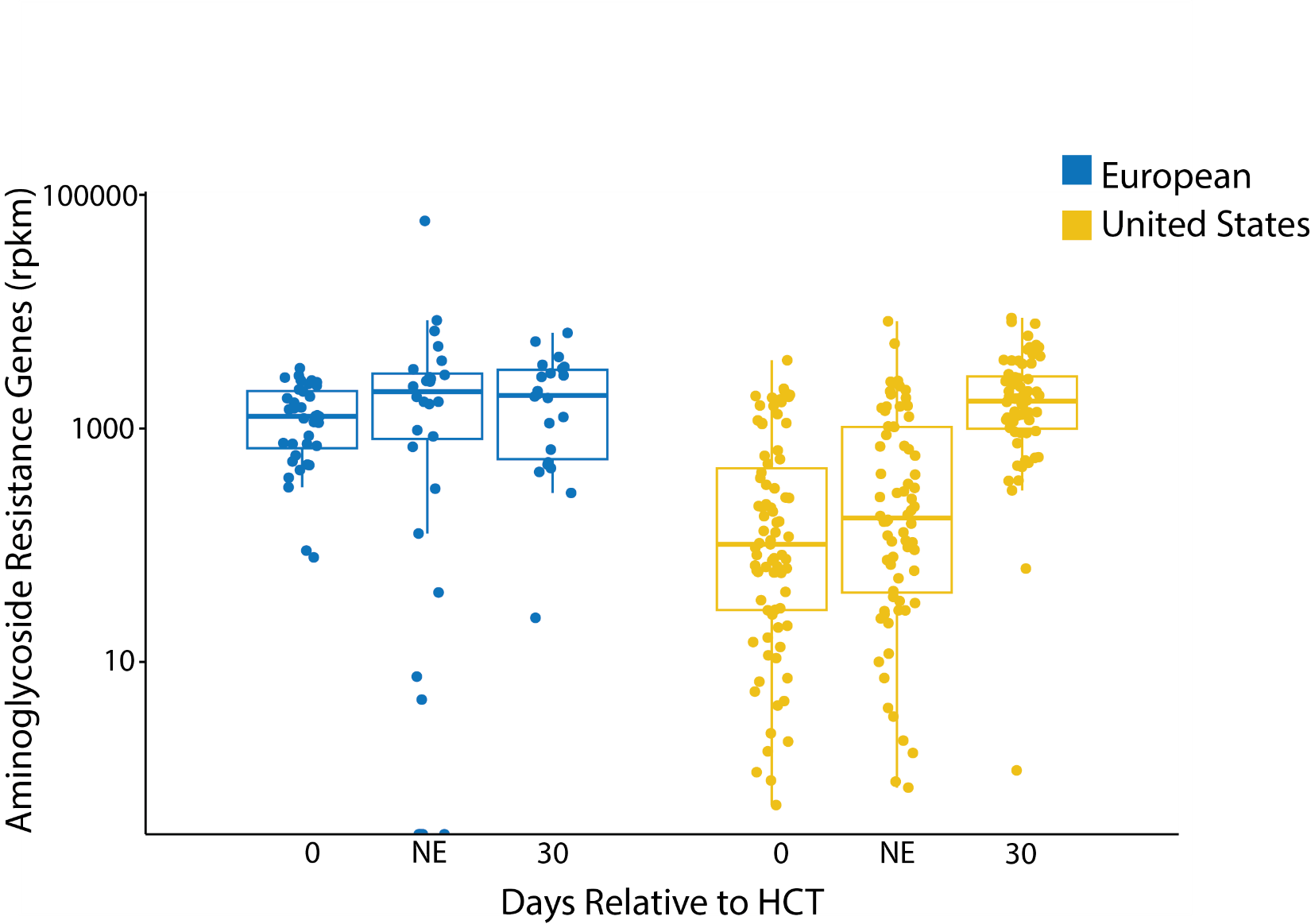

**Supplemental Figure 4: Abundance of ARGs associated with aminoglycoside resistance.** Antibiotic resistance genes conferring resistance to aminoglycosides do not follow a similar pattern to ARGs conferring ESBL resistance to beta-lactams

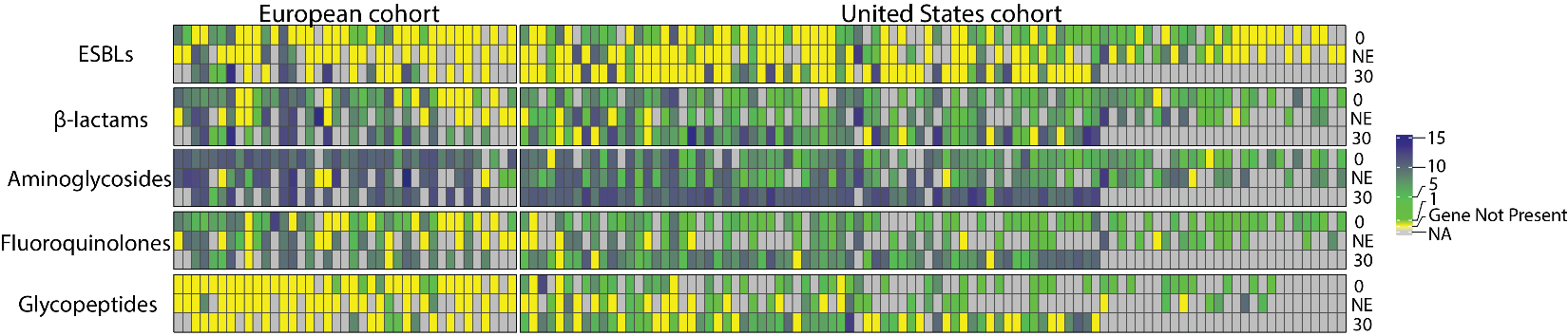

**supplemental Figure 5: ARG abundance heatmap based on antibiotic class.** “0” refers to the sample collected at HCT, “NE” refers to the sample collected at neutrophil engraftment, and “30” refers to the sample collected 30 days after HCT. Each individual is one column, with the abundance of antibiotic resistance gene represented by a color.

**Supplemental Tables:**

**Supplemental table 1: Antibiotics Received between day 0 and day 30**. The United States cohort saw double the average daily doses of antibiotics in the study period primarily because the St. Jude HCT protocol calls for daily prophylactic antibiotics. All patients in the United States cohort received daily trimethoprim-sulfamethoxazole prophylaxis (unless contra-indicated then inhaled pentamidine was administered). Starting in 2020 high-risk participants received fluoroquinolone prophylaxis during neutropenia. The European cohort protocol did not call for prophylactic antibiotics during neutropenia.

|  | European Cohort (N = 39) | United States Cohort (N = 94) | p-value |
| --- | --- | --- | --- |
| Days on aminoglycosides, Median [Min, Max] | 0.0 [0.0, 19.0] | 0.0 [0.0, 15.0] | <0.001 |
| Aminoglycoside exposure, n (%) | 18 (46.2%) | 8 (8.5%) | <0.001 |
| Days on beta-lactams, Median [Min, Max] | 14.0 [0.0, 41.0] | 9.5 [0.0, 62.0] | 0.017 |
| Beta-lactams exposure, n (%) | 33 (84.6%) | 89 (94.7%) | 0.081 |
| Days on fluoroquinolones, Median [Min, Max] | 0.0 [0.0, 16.0] | 1.0 [0.0, 63.0] | <0.001 |
| Fluoroquinolone exposure, n (%) | 3 (7.7%) | 48 (51.1%) | <0.001 |
| Days on glycopeptides, Median [Min, Max] | 0.0 [0.0, 23.0] | 4.0 [0.0, 41.0] | 0.002 |
| Glycopeptide exposure, n (%) | 10 (25.6%) | 70 (74.5%) | <0.001 |
| Days on anaerobic (active against *Bacteroides fragilis*), Median [Min, Max] | 9.0 [0.0, 31.0] | 0.0 [0.0, 47.0] | 0.001 |
| Anaerobic (active against *Bacteroides fragilis*), exposure, n (%) | 26 (66.7%) | 40 (42.6%) | 0.011 |
| Days on ESBL-susceptible beta-lactams, Median [Min, Max] | 0.0 [0.0, 21.0] | 8.0 [0.0, 22.0] | <0.001 |
| ESBL-susceptible beta-lactam exposure, n (%) | 8 (20.5%) | 89 (94.7%) | <0.001 |
| Total days on antibiotics, Median [Min, Max] | 25.0 [0.0, 58.0] | 56.0 [1.0, 146.0] | <0.001 |
| Any antibiotic exposure, n (%) | 34 (87.2%) | 94 (100.0%) | 0.002 |

**Supplemental table 2: Associations with antibiotic exposures and ESBL gene accumulation before neutrophil engraftment.** Only aminoglycosides are associated with increases in ESBL genes detected at neutrophil engraftment for the European cohort. Aminoglycosides were too infrequently administered in the United States cohort to be compared. For both cohorts, total antibiotic doses or daily doses of beta-lactams, or beta-lactams cleavable by ESBL enzymes, or all antibiotics with anaerobic activity did not correlate with changes in ESBL abundance. P-values adjusted for multiple comparisons and confounding variables using CBPS weighting.

|  | **European Cohort** | | | | **United States Cohort** | | | |
| --- | --- | --- | --- | --- | --- | --- | --- | --- |
|  | **Univariate** | | **CBPS weighted** | | **Univariate** | | **CBPS weighted** | |
| **Antibiotic exposure** | **Est. (95% CI)** | **P** | **Est. (95% CI)** | **P** | **Est. (95% CI)** | **P** | **Est. (95% CI)** | **P** |
| Beta-lactams | 0.18 [-0.15; 0.51] | 0.27 | 0.071 [-0.29; 0.43] | 0.685 | -0.11 [-0.35; 0.13] | 0.355 | -0.077 [-0.24; 0.088] | 0.353 |
| Aminoglycosides | 1.30 [0.64; 1.95] | **<0.001** | 0.93 [0.38; 1.49] | **0.002** |  |  |  |  |
| beta-lactams susceptible to ESBLs |  |  |  |  | 0.0017 [-0.21; 0.22] | 0.988 | -0.0088 [-0.22; 0.2] | 0.933 |
| Anaerobic coverage (active against *Bacteroides fragilis*) | 0.21 [-0.097; 0.53] | 0.167 | 0.038 [-0.53; 0.61] | 0.893 | -0.18 [-0.53; 0.17] | 0.300 | -0.38 [-0.8; 0.036] | 0.073 |
| Total antibiotic (daily doses) | 0.039 [-0.16; 0.24] | 0.691 | -0.057 [-0.31; 0.19] | 0.643 | -0.031 [-0.067; 0.0047] | 0.087 | -0.0071 [-0.067; 0.053] | 0.815 |

**Supplemental table 3: Associations with antibiotic exposures and ESBL gene accumulation in the first 30 days post HCT.** For both cohorts, total antibiotic doses or daily doses of beta-lactams, or beta-lactams cleavable by ESBL enzymes, aminoglycosides, or all antibiotics with anaerobic activity did not correlate with changes in ESBL abundance. P-values adjusted for multiple comparisons and confounding variables using CBPS weighting.

|  | **European Cohort** | | | | **United States Cohort** | | | |
| --- | --- | --- | --- | --- | --- | --- | --- | --- |
|  | **Univariate** | | **CBPS weighted** | | **Univariate** | | **CBPS weighted** | |
| **Antibiotic exposure** | **Est. (95% CI)** | **P** | **Est. (95% CI)** | **P** | **Est. (95% CI)** | **P** | **Est. (95% CI)** | **P** |
| Beta-lactams | 0.091 [-0.21; 0.39] | 0.536 | 0.034 [-0.34; 0.4] | 0.849 | -0.1 [-0.96; 0.75] | 0.811 | -0.056 [-0.28; 0.17] | 0.623 |
| Aminoglycosides | 0.024 [-2.34; 2.39] | 0.983 | -0.025 [-2.55; 2.50] | 0.984 |  |  |  |  |
| beta-lactams susceptible to ESBLs |  |  |  |  | -0.24 [-0.48; 0.0093] | 0.059 | -0.48 [-1.04; 0.066] | 0.083 |
| Anaerobic coverage (active against *Bacteroides fragilis*) | 0.18 [-0.16; 0.52] | 0.276 | -0.04 [-0.47; 0.39] | 0.848 | 0.017 [-0.2; 0.24] | 0.877 | 0.027 [-0.21; 0.26] | 0.818 |
| total | 0.0094 [-0.2; 0.22] | 0.925 | -0.049 [-0.28; 0.18] | 0.661 | -0.011 [-0.092; 0.069] | 0.779 | 0.0057 [-0.073; 0.085] | 0.886 |

**Supplemental Table 4: Pre-transplant resistome disruption predicts ESBL gene accumulation during early transplant.** Bray-Curtis distance from the age-matched healthy-cohort centroid of the resistome at baseline is associated with increased ESBL gene abundance at subsequent timepoints (neutrophil engraftment and day 30). Associations were assessed using univariate logistic regression with propensity score weighting.

|  |  | **European cohort** | | **United States cohort** | |
| --- | --- | --- | --- | --- | --- |
| **Association** | **Time period** | **Est. (SE)** | **p** | **Est. (SE)** | **p** |
| Resistome Disturbance (Bray-Curtis distance from Healthy Centroid) | Baseline to neutrophil engraftment | 7.48 (3.65) | 0.052 | **15.85 (6.19)** | **0.024** |
| Resistome Disturbance (Bray-Curtis distance from Healthy Centroid) | First 30 days | **15.12 (5.51)** | **0.013** | 9.58 (4.99) | 0.079 |

**Supplemental table 5: Genes encoding ESBLs in the MEGARes database.** Genes conferring extended spectrum beta-lactamase enzymes were identified by characterization on the Beta-lactamase Database maintained by Bush and Jacoby^4^.

| MEGARes header | Gene | MDRO phenotype |
| --- | --- | --- |
| MEG_3107\|Drugs\|betalactams\|Class_A_betalactamases\|GES | GES | ESBL |
| MEG_3108\|Drugs\|betalactams\|Class_A_betalactamases\|GES | GES | ESBL |
| MEG_3109\|Drugs\|betalactams\|Class_A_betalactamases\|GES | GES | ESBL |
| MEG_3110\|Drugs\|betalactams\|Class_A_betalactamases\|GES | GES | ESBL |
| MEG_3112\|Drugs\|betalactams\|Class_A_betalactamases\|GES | GES | ESBL |
| MEG_3113\|Drugs\|betalactams\|Class_A_betalactamases\|GES | GES | ESBL |
| MEG_3114\|Drugs\|betalactams\|Class_A_betalactamases\|GES | GES | ESBL |
| MEG_3115\|Drugs\|betalactams\|Class_A_betalactamases\|GES | GES | ESBL |
| MEG_3116\|Drugs\|betalactams\|Class_A_betalactamases\|GES | GES | ESBL |
| MEG_3118\|Drugs\|betalactams\|Class_A_betalactamases\|GES | GES | ESBL |
| MEG_3119\|Drugs\|betalactams\|Class_A_betalactamases\|GES | GES | ESBL |
| MEG_3120\|Drugs\|betalactams\|Class_A_betalactamases\|GES | GES | ESBL |
| MEG_3124\|Drugs\|betalactams\|Class_A_betalactamases\|GES | GES | ESBL |
| MEG_3128\|Drugs\|betalactams\|Class_A_betalactamases\|GES | GES | ESBL |
| MEG_3106\|Drugs\|betalactams\|Class_A_betalactamases\|GES | GES | ESBL |
| MEG_3085\|Drugs\|betalactams\|Class_A_betalactamases\|GES | GES | ESBL |
| MEG_3104\|Drugs\|betalactams\|Class_A_betalactamases\|GES | GES | ESBL |
| MEG_3114\|Drugs\|betalactams\|Class_A_betalactamases\|GES | GES | ESBL |
| MEG_3117\|Drugs\|betalactams\|Class_A_betalactamases\|GES | GES | ESBL |
| MEG_3121\|Drugs\|betalactams\|Class_A_betalactamases\|GES | GES | ESBL |
| MEG_3128\|Drugs\|betalactams\|Class_A_betalactamases\|GES | GES | ESBL |
| MEG_8038\|Drugs\|betalactams\|Class_A_betalactamases\|CTX | CTX | ESBL |
| MEG_8039\|Drugs\|betalactams\|Class_A_betalactamases\|CTX | CTX | ESBL |
| MEG_8040\|Drugs\|betalactams\|Class_A_betalactamases\|CTX | CTX | ESBL |
| MEG_8041\|Drugs\|betalactams\|Class_A_betalactamases\|CTX | CTX | ESBL |
| MEG_8042\|Drugs\|betalactams\|Class_A_betalactamases\|CTX | CTX | ESBL |
| MEG_8043\|Drugs\|betalactams\|Class_A_betalactamases\|CTX | CTX | ESBL |
| MEG_8044\|Drugs\|betalactams\|Class_A_betalactamases\|CTX | CTX | ESBL |
| MEG_8045\|Drugs\|betalactams\|Class_A_betalactamases\|CTX | CTX | ESBL |
| MEG_8046\|Drugs\|betalactams\|Class_A_betalactamases\|CTX | CTX | ESBL |
| MEG_8047\|Drugs\|betalactams\|Class_A_betalactamases\|CTX | CTX | ESBL |
| MEG_8048\|Drugs\|betalactams\|Class_A_betalactamases\|CTX | CTX | ESBL |
| MEG_8049\|Drugs\|betalactams\|Class_A_betalactamases\|CTX | CTX | ESBL |
| MEG_8050\|Drugs\|betalactams\|Class_A_betalactamases\|CTX | CTX | ESBL |
| MEG_8051\|Drugs\|betalactams\|Class_A_betalactamases\|CTX | CTX | ESBL |
| MEG_8052\|Drugs\|betalactams\|Class_A_betalactamases\|CTX | CTX | ESBL |
| MEG_8053\|Drugs\|betalactams\|Class_A_betalactamases\|CTX | CTX | ESBL |
| MEG_8054\|Drugs\|betalactams\|Class_A_betalactamases\|CTX | CTX | ESBL |
| MEG_8055\|Drugs\|betalactams\|Class_A_betalactamases\|CTX | CTX | ESBL |
| MEG_8056\|Drugs\|betalactams\|Class_A_betalactamases\|CTX | CTX | ESBL |
| MEG_8057\|Drugs\|betalactams\|Class_A_betalactamases\|CTX | CTX | ESBL |
| MEG_8058\|Drugs\|betalactams\|Class_A_betalactamases\|CTX | CTX | ESBL |
| MEG_8059\|Drugs\|betalactams\|Class_A_betalactamases\|CTX | CTX | ESBL |
| MEG_2144\|Drugs\|betalactams\|Class_A_betalactamases\|CTX | CTX | ESBL |
| MEG_2145\|Drugs\|betalactams\|Class_A_betalactamases\|CTX | CTX | ESBL |
| MEG_2146\|Drugs\|betalactams\|Class_A_betalactamases\|CTX | CTX | ESBL |
| MEG_2147\|Drugs\|betalactams\|Class_A_betalactamases\|CTX | CTX | ESBL |
| MEG_2148\|Drugs\|betalactams\|Class_A_betalactamases\|CTX | CTX | ESBL |
| MEG_2149\|Drugs\|betalactams\|Class_A_betalactamases\|CTX | CTX | ESBL |
| MEG_2150\|Drugs\|betalactams\|Class_A_betalactamases\|CTX | CTX | ESBL |
| MEG_2151\|Drugs\|betalactams\|Class_A_betalactamases\|CTX | CTX | ESBL |
| MEG_2152\|Drugs\|betalactams\|Class_A_betalactamases\|CTX | CTX | ESBL |
| MEG_2153\|Drugs\|betalactams\|Class_A_betalactamases\|CTX | CTX | ESBL |
| MEG_2154\|Drugs\|betalactams\|Class_A_betalactamases\|CTX | CTX | ESBL |
| MEG_2155\|Drugs\|betalactams\|Class_A_betalactamases\|CTX | CTX | ESBL |
| MEG_2156\|Drugs\|betalactams\|Class_A_betalactamases\|CTX | CTX | ESBL |
| MEG_2157\|Drugs\|betalactams\|Class_A_betalactamases\|CTX | CTX | ESBL |
| MEG_2158\|Drugs\|betalactams\|Class_A_betalactamases\|CTX | CTX | ESBL |
| MEG_2159\|Drugs\|betalactams\|Class_A_betalactamases\|CTX | CTX | ESBL |
| MEG_2160\|Drugs\|betalactams\|Class_A_betalactamases\|CTX | CTX | ESBL |
| MEG_2161\|Drugs\|betalactams\|Class_A_betalactamases\|CTX | CTX | ESBL |
| MEG_2162\|Drugs\|betalactams\|Class_A_betalactamases\|CTX | CTX | ESBL |
| MEG_2163\|Drugs\|betalactams\|Class_A_betalactamases\|CTX | CTX | ESBL |
| MEG_2164\|Drugs\|betalactams\|Class_A_betalactamases\|CTX | CTX | ESBL |
| MEG_2165\|Drugs\|betalactams\|Class_A_betalactamases\|CTX | CTX | ESBL |
| MEG_2166\|Drugs\|betalactams\|Class_A_betalactamases\|CTX | CTX | ESBL |
| MEG_2167\|Drugs\|betalactams\|Class_A_betalactamases\|CTX | CTX | ESBL |
| MEG_2168\|Drugs\|betalactams\|Class_A_betalactamases\|CTX | CTX | ESBL |
| MEG_2169\|Drugs\|betalactams\|Class_A_betalactamases\|CTX | CTX | ESBL |
| MEG_2170\|Drugs\|betalactams\|Class_A_betalactamases\|CTX | CTX | ESBL |
| MEG_2171\|Drugs\|betalactams\|Class_A_betalactamases\|CTX | CTX | ESBL |
| MEG_2172\|Drugs\|betalactams\|Class_A_betalactamases\|CTX | CTX | ESBL |
| MEG_2173\|Drugs\|betalactams\|Class_A_betalactamases\|CTX | CTX | ESBL |
| MEG_2174\|Drugs\|betalactams\|Class_A_betalactamases\|CTX | CTX | ESBL |
| MEG_2175\|Drugs\|betalactams\|Class_A_betalactamases\|CTX | CTX | ESBL |
| MEG_2176\|Drugs\|betalactams\|Class_A_betalactamases\|CTX | CTX | ESBL |
| MEG_2177\|Drugs\|betalactams\|Class_A_betalactamases\|CTX | CTX | ESBL |
| MEG_2178\|Drugs\|betalactams\|Class_A_betalactamases\|CTX | CTX | ESBL |
| MEG_2179\|Drugs\|betalactams\|Class_A_betalactamases\|CTX | CTX | ESBL |
| MEG_2180\|Drugs\|betalactams\|Class_A_betalactamases\|CTX | CTX | ESBL |
| MEG_2181\|Drugs\|betalactams\|Class_A_betalactamases\|CTX | CTX | ESBL |
| MEG_2182\|Drugs\|betalactams\|Class_A_betalactamases\|CTX | CTX | ESBL |
| MEG_2183\|Drugs\|betalactams\|Class_A_betalactamases\|CTX | CTX | ESBL |
| MEG_2184\|Drugs\|betalactams\|Class_A_betalactamases\|CTX | CTX | ESBL |
| MEG_2185\|Drugs\|betalactams\|Class_A_betalactamases\|CTX | CTX | ESBL |
| MEG_2186\|Drugs\|betalactams\|Class_A_betalactamases\|CTX | CTX | ESBL |
| MEG_2187\|Drugs\|betalactams\|Class_A_betalactamases\|CTX | CTX | ESBL |
| MEG_2188\|Drugs\|betalactams\|Class_A_betalactamases\|CTX | CTX | ESBL |
| MEG_2189\|Drugs\|betalactams\|Class_A_betalactamases\|CTX | CTX | ESBL |
| MEG_2190\|Drugs\|betalactams\|Class_A_betalactamases\|CTX | CTX | ESBL |
| MEG_2191\|Drugs\|betalactams\|Class_A_betalactamases\|CTX | CTX | ESBL |
| MEG_2192\|Drugs\|betalactams\|Class_A_betalactamases\|CTX | CTX | ESBL |
| MEG_2193\|Drugs\|betalactams\|Class_A_betalactamases\|CTX | CTX | ESBL |
| MEG_2194\|Drugs\|betalactams\|Class_A_betalactamases\|CTX | CTX | ESBL |
| MEG_2195\|Drugs\|betalactams\|Class_A_betalactamases\|CTX | CTX | ESBL |
| MEG_2196\|Drugs\|betalactams\|Class_A_betalactamases\|CTX | CTX | ESBL |
| MEG_2197\|Drugs\|betalactams\|Class_A_betalactamases\|CTX | CTX | ESBL |
| MEG_2198\|Drugs\|betalactams\|Class_A_betalactamases\|CTX | CTX | ESBL |
| MEG_2199\|Drugs\|betalactams\|Class_A_betalactamases\|CTX | CTX | ESBL |
| MEG_2200\|Drugs\|betalactams\|Class_A_betalactamases\|CTX | CTX | ESBL |
| MEG_2201\|Drugs\|betalactams\|Class_A_betalactamases\|CTX | CTX | ESBL |
| MEG_2202\|Drugs\|betalactams\|Class_A_betalactamases\|CTX | CTX | ESBL |
| MEG_2203\|Drugs\|betalactams\|Class_A_betalactamases\|CTX | CTX | ESBL |
| MEG_2204\|Drugs\|betalactams\|Class_A_betalactamases\|CTX | CTX | ESBL |
| MEG_2205\|Drugs\|betalactams\|Class_A_betalactamases\|CTX | CTX | ESBL |
| MEG_2206\|Drugs\|betalactams\|Class_A_betalactamases\|CTX | CTX | ESBL |
| MEG_2207\|Drugs\|betalactams\|Class_A_betalactamases\|CTX | CTX | ESBL |
| MEG_2208\|Drugs\|betalactams\|Class_A_betalactamases\|CTX | CTX | ESBL |
| MEG_2209\|Drugs\|betalactams\|Class_A_betalactamases\|CTX | CTX | ESBL |
| MEG_2210\|Drugs\|betalactams\|Class_A_betalactamases\|CTX | CTX | ESBL |
| MEG_2211\|Drugs\|betalactams\|Class_A_betalactamases\|CTX | CTX | ESBL |
| MEG_2212\|Drugs\|betalactams\|Class_A_betalactamases\|CTX | CTX | ESBL |
| MEG_2213\|Drugs\|betalactams\|Class_A_betalactamases\|CTX | CTX | ESBL |
| MEG_2214\|Drugs\|betalactams\|Class_A_betalactamases\|CTX | CTX | ESBL |
| MEG_2215\|Drugs\|betalactams\|Class_A_betalactamases\|CTX | CTX | ESBL |
| MEG_2216\|Drugs\|betalactams\|Class_A_betalactamases\|CTX | CTX | ESBL |
| MEG_2217\|Drugs\|betalactams\|Class_A_betalactamases\|CTX | CTX | ESBL |
| MEG_2218\|Drugs\|betalactams\|Class_A_betalactamases\|CTX | CTX | ESBL |
| MEG_2219\|Drugs\|betalactams\|Class_A_betalactamases\|CTX | CTX | ESBL |
| MEG_2220\|Drugs\|betalactams\|Class_A_betalactamases\|CTX | CTX | ESBL |
| MEG_2221\|Drugs\|betalactams\|Class_A_betalactamases\|CTX | CTX | ESBL |
| MEG_2222\|Drugs\|betalactams\|Class_A_betalactamases\|CTX | CTX | ESBL |
| MEG_2223\|Drugs\|betalactams\|Class_A_betalactamases\|CTX | CTX | ESBL |
| MEG_2224\|Drugs\|betalactams\|Class_A_betalactamases\|CTX | CTX | ESBL |
| MEG_2225\|Drugs\|betalactams\|Class_A_betalactamases\|CTX | CTX | ESBL |
| MEG_2226\|Drugs\|betalactams\|Class_A_betalactamases\|CTX | CTX | ESBL |
| MEG_2227\|Drugs\|betalactams\|Class_A_betalactamases\|CTX | CTX | ESBL |
| MEG_2228\|Drugs\|betalactams\|Class_A_betalactamases\|CTX | CTX | ESBL |
| MEG_2229\|Drugs\|betalactams\|Class_A_betalactamases\|CTX | CTX | ESBL |
| MEG_2230\|Drugs\|betalactams\|Class_A_betalactamases\|CTX | CTX | ESBL |
| MEG_2231\|Drugs\|betalactams\|Class_A_betalactamases\|CTX | CTX | ESBL |
| MEG_2232\|Drugs\|betalactams\|Class_A_betalactamases\|CTX | CTX | ESBL |
| MEG_2233\|Drugs\|betalactams\|Class_A_betalactamases\|CTX | CTX | ESBL |
| MEG_2234\|Drugs\|betalactams\|Class_A_betalactamases\|CTX | CTX | ESBL |
| MEG_2235\|Drugs\|betalactams\|Class_A_betalactamases\|CTX | CTX | ESBL |
| MEG_2236\|Drugs\|betalactams\|Class_A_betalactamases\|CTX | CTX | ESBL |
| MEG_2237\|Drugs\|betalactams\|Class_A_betalactamases\|CTX | CTX | ESBL |
| MEG_2238\|Drugs\|betalactams\|Class_A_betalactamases\|CTX | CTX | ESBL |
| MEG_2239\|Drugs\|betalactams\|Class_A_betalactamases\|CTX | CTX | ESBL |
| MEG_2240\|Drugs\|betalactams\|Class_A_betalactamases\|CTX | CTX | ESBL |
| MEG_2241\|Drugs\|betalactams\|Class_A_betalactamases\|CTX | CTX | ESBL |
| MEG_2242\|Drugs\|betalactams\|Class_A_betalactamases\|CTX | CTX | ESBL |
| MEG_2243\|Drugs\|betalactams\|Class_A_betalactamases\|CTX | CTX | ESBL |
| MEG_2244\|Drugs\|betalactams\|Class_A_betalactamases\|CTX | CTX | ESBL |
| MEG_2245\|Drugs\|betalactams\|Class_A_betalactamases\|CTX | CTX | ESBL |
| MEG_2246\|Drugs\|betalactams\|Class_A_betalactamases\|CTX | CTX | ESBL |
| MEG_2247\|Drugs\|betalactams\|Class_A_betalactamases\|CTX | CTX | ESBL |
| MEG_2248\|Drugs\|betalactams\|Class_A_betalactamases\|CTX | CTX | ESBL |
| MEG_2249\|Drugs\|betalactams\|Class_A_betalactamases\|CTX | CTX | ESBL |
| MEG_2250\|Drugs\|betalactams\|Class_A_betalactamases\|CTX | CTX | ESBL |
| MEG_2251\|Drugs\|betalactams\|Class_A_betalactamases\|CTX | CTX | ESBL |
| MEG_2252\|Drugs\|betalactams\|Class_A_betalactamases\|CTX | CTX | ESBL |
| MEG_2253\|Drugs\|betalactams\|Class_A_betalactamases\|CTX | CTX | ESBL |
| MEG_2254\|Drugs\|betalactams\|Class_A_betalactamases\|CTX | CTX | ESBL |
| MEG_2255\|Drugs\|betalactams\|Class_A_betalactamases\|CTX | CTX | ESBL |
| MEG_2256\|Drugs\|betalactams\|Class_A_betalactamases\|CTX | CTX | ESBL |
| MEG_2257\|Drugs\|betalactams\|Class_A_betalactamases\|CTX | CTX | ESBL |
| MEG_2258\|Drugs\|betalactams\|Class_A_betalactamases\|CTX | CTX | ESBL |
| MEG_2259\|Drugs\|betalactams\|Class_A_betalactamases\|CTX | CTX | ESBL |
| MEG_2260\|Drugs\|betalactams\|Class_A_betalactamases\|CTX | CTX | ESBL |
| MEG_2261\|Drugs\|betalactams\|Class_A_betalactamases\|CTX | CTX | ESBL |
| MEG_2262\|Drugs\|betalactams\|Class_A_betalactamases\|CTX | CTX | ESBL |
| MEG_2263\|Drugs\|betalactams\|Class_A_betalactamases\|CTX | CTX | ESBL |
| MEG_2264\|Drugs\|betalactams\|Class_A_betalactamases\|CTX | CTX | ESBL |
| MEG_2265\|Drugs\|betalactams\|Class_A_betalactamases\|CTX | CTX | ESBL |
| MEG_2266\|Drugs\|betalactams\|Class_A_betalactamases\|CTX | CTX | ESBL |
| MEG_2267\|Drugs\|betalactams\|Class_A_betalactamases\|CTX | CTX | ESBL |
| MEG_2268\|Drugs\|betalactams\|Class_A_betalactamases\|CTX | CTX | ESBL |
| MEG_2269\|Drugs\|betalactams\|Class_A_betalactamases\|CTX | CTX | ESBL |
| MEG_2270\|Drugs\|betalactams\|Class_A_betalactamases\|CTX | CTX | ESBL |
| MEG_2271\|Drugs\|betalactams\|Class_A_betalactamases\|CTX | CTX | ESBL |
| MEG_2272\|Drugs\|betalactams\|Class_A_betalactamases\|CTX | CTX | ESBL |
| MEG_2273\|Drugs\|betalactams\|Class_A_betalactamases\|CTX | CTX | ESBL |
| MEG_2274\|Drugs\|betalactams\|Class_A_betalactamases\|CTX | CTX | ESBL |
| MEG_2275\|Drugs\|betalactams\|Class_A_betalactamases\|CTX | CTX | ESBL |
| MEG_2276\|Drugs\|betalactams\|Class_A_betalactamases\|CTX | CTX | ESBL |
| MEG_2277\|Drugs\|betalactams\|Class_A_betalactamases\|CTX | CTX | ESBL |
| MEG_2278\|Drugs\|betalactams\|Class_A_betalactamases\|CTX | CTX | ESBL |
| MEG_2279\|Drugs\|betalactams\|Class_A_betalactamases\|CTX | CTX | ESBL |
| MEG_2280\|Drugs\|betalactams\|Class_A_betalactamases\|CTX | CTX | ESBL |
| MEG_2281\|Drugs\|betalactams\|Class_A_betalactamases\|CTX | CTX | ESBL |
| MEG_2282\|Drugs\|betalactams\|Class_A_betalactamases\|CTX | CTX | ESBL |
| MEG_2283\|Drugs\|betalactams\|Class_A_betalactamases\|CTX | CTX | ESBL |
| MEG_2284\|Drugs\|betalactams\|Class_A_betalactamases\|CTX | CTX | ESBL |
| MEG_2285\|Drugs\|betalactams\|Class_A_betalactamases\|CTX | CTX | ESBL |
| MEG_2286\|Drugs\|betalactams\|Class_A_betalactamases\|CTX | CTX | ESBL |
| MEG_2287\|Drugs\|betalactams\|Class_A_betalactamases\|CTX | CTX | ESBL |
| MEG_2288\|Drugs\|betalactams\|Class_A_betalactamases\|CTX | CTX | ESBL |
| MEG_2289\|Drugs\|betalactams\|Class_A_betalactamases\|CTX | CTX | ESBL |
| MEG_2290\|Drugs\|betalactams\|Class_A_betalactamases\|CTX | CTX | ESBL |
| MEG_2291\|Drugs\|betalactams\|Class_A_betalactamases\|CTX | CTX | ESBL |
| MEG_2292\|Drugs\|betalactams\|Class_A_betalactamases\|CTX | CTX | ESBL |
| MEG_2293\|Drugs\|betalactams\|Class_A_betalactamases\|CTX | CTX | ESBL |
| MEG_2294\|Drugs\|betalactams\|Class_A_betalactamases\|CTX | CTX | ESBL |
| MEG_2295\|Drugs\|betalactams\|Class_A_betalactamases\|CTX | CTX | ESBL |
| MEG_2296\|Drugs\|betalactams\|Class_A_betalactamases\|CTX | CTX | ESBL |
| MEG_2297\|Drugs\|betalactams\|Class_A_betalactamases\|CTX | CTX | ESBL |
| MEG_2298\|Drugs\|betalactams\|Class_A_betalactamases\|CTX | CTX | ESBL |
| MEG_2299\|Drugs\|betalactams\|Class_A_betalactamases\|CTX | CTX | ESBL |
| MEG_2300\|Drugs\|betalactams\|Class_A_betalactamases\|CTX | CTX | ESBL |
| MEG_2301\|Drugs\|betalactams\|Class_A_betalactamases\|CTX | CTX | ESBL |
| MEG_2302\|Drugs\|betalactams\|Class_A_betalactamases\|CTX | CTX | ESBL |
| MEG_2303\|Drugs\|betalactams\|Class_A_betalactamases\|CTX | CTX | ESBL |
| MEG_2304\|Drugs\|betalactams\|Class_A_betalactamases\|CTX | CTX | ESBL |
| MEG_2305\|Drugs\|betalactams\|Class_A_betalactamases\|CTX | CTX | ESBL |
| MEG_2306\|Drugs\|betalactams\|Class_A_betalactamases\|CTX | CTX | ESBL |
| MEG_2307\|Drugs\|betalactams\|Class_A_betalactamases\|CTX | CTX | ESBL |
| MEG_2308\|Drugs\|betalactams\|Class_A_betalactamases\|CTX | CTX | ESBL |
| MEG_2309\|Drugs\|betalactams\|Class_A_betalactamases\|CTX | CTX | ESBL |
| MEG_2310\|Drugs\|betalactams\|Class_A_betalactamases\|CTX | CTX | ESBL |
| MEG_2311\|Drugs\|betalactams\|Class_A_betalactamases\|CTX | CTX | ESBL |
| MEG_2312\|Drugs\|betalactams\|Class_A_betalactamases\|CTX | CTX | ESBL |
| MEG_2313\|Drugs\|betalactams\|Class_A_betalactamases\|CTX | CTX | ESBL |
| MEG_2314\|Drugs\|betalactams\|Class_A_betalactamases\|CTX | CTX | ESBL |
| MEG_2315\|Drugs\|betalactams\|Class_A_betalactamases\|CTX | CTX | ESBL |
| MEG_2316\|Drugs\|betalactams\|Class_A_betalactamases\|CTX | CTX | ESBL |
| MEG_2317\|Drugs\|betalactams\|Class_A_betalactamases\|CTX | CTX | ESBL |
| MEG_2318\|Drugs\|betalactams\|Class_A_betalactamases\|CTX | CTX | ESBL |
| MEG_2319\|Drugs\|betalactams\|Class_A_betalactamases\|CTX | CTX | ESBL |
| MEG_2320\|Drugs\|betalactams\|Class_A_betalactamases\|CTX | CTX | ESBL |
| MEG_2321\|Drugs\|betalactams\|Class_A_betalactamases\|CTX | CTX | ESBL |
| MEG_2322\|Drugs\|betalactams\|Class_A_betalactamases\|CTX | CTX | ESBL |
| MEG_2323\|Drugs\|betalactams\|Class_A_betalactamases\|CTX | CTX | ESBL |
| MEG_2324\|Drugs\|betalactams\|Class_A_betalactamases\|CTX | CTX | ESBL |
| MEG_2325\|Drugs\|betalactams\|Class_A_betalactamases\|CTX | CTX | ESBL |
| MEG_2326\|Drugs\|betalactams\|Class_A_betalactamases\|CTX | CTX | ESBL |
| MEG_2327\|Drugs\|betalactams\|Class_A_betalactamases\|CTX | CTX | ESBL |
| MEG_2328\|Drugs\|betalactams\|Class_A_betalactamases\|CTX | CTX | ESBL |
| MEG_2329\|Drugs\|betalactams\|Class_A_betalactamases\|CTX | CTX | ESBL |
| MEG_2330\|Drugs\|betalactams\|Class_A_betalactamases\|CTX | CTX | ESBL |
| MEG_2331\|Drugs\|betalactams\|Class_A_betalactamases\|CTX | CTX | ESBL |
| MEG_2332\|Drugs\|betalactams\|Class_A_betalactamases\|CTX | CTX | ESBL |
| MEG_2333\|Drugs\|betalactams\|Class_A_betalactamases\|CTX | CTX | ESBL |
| MEG_2334\|Drugs\|betalactams\|Class_A_betalactamases\|CTX | CTX | ESBL |
| MEG_2335\|Drugs\|betalactams\|Class_A_betalactamases\|CTX | CTX | ESBL |
| MEG_2336\|Drugs\|betalactams\|Class_A_betalactamases\|CTX | CTX | ESBL |
| MEG_2337\|Drugs\|betalactams\|Class_A_betalactamases\|CTX | CTX | ESBL |
| MEG_2338\|Drugs\|betalactams\|Class_A_betalactamases\|CTX | CTX | ESBL |
| MEG_2339\|Drugs\|betalactams\|Class_A_betalactamases\|CTX | CTX | ESBL |
| MEG_2340\|Drugs\|betalactams\|Class_A_betalactamases\|CTX | CTX | ESBL |
| MEG_2341\|Drugs\|betalactams\|Class_A_betalactamases\|CTX | CTX | ESBL |
| MEG_2342\|Drugs\|betalactams\|Class_A_betalactamases\|CTX | CTX | ESBL |
| MEG_2343\|Drugs\|betalactams\|Class_A_betalactamases\|CTX | CTX | ESBL |
| MEG_2344\|Drugs\|betalactams\|Class_A_betalactamases\|CTX | CTX | ESBL |
| MEG_2345\|Drugs\|betalactams\|Class_A_betalactamases\|CTX | CTX | ESBL |
| MEG_2346\|Drugs\|betalactams\|Class_A_betalactamases\|CTX | CTX | ESBL |
| MEG_2347\|Drugs\|betalactams\|Class_A_betalactamases\|CTX | CTX | ESBL |
| MEG_2348\|Drugs\|betalactams\|Class_A_betalactamases\|CTX | CTX | ESBL |
| MEG_2349\|Drugs\|betalactams\|Class_A_betalactamases\|CTX | CTX | ESBL |
| MEG_2350\|Drugs\|betalactams\|Class_A_betalactamases\|CTX | CTX | ESBL |
| MEG_2351\|Drugs\|betalactams\|Class_A_betalactamases\|CTX | CTX | ESBL |
| MEG_2352\|Drugs\|betalactams\|Class_A_betalactamases\|CTX | CTX | ESBL |
| MEG_2353\|Drugs\|betalactams\|Class_A_betalactamases\|CTX | CTX | ESBL |
| MEG_2354\|Drugs\|betalactams\|Class_A_betalactamases\|CTX | CTX | ESBL |
| MEG_2355\|Drugs\|betalactams\|Class_A_betalactamases\|CTX | CTX | ESBL |
| MEG_2356\|Drugs\|betalactams\|Class_A_betalactamases\|CTX | CTX | ESBL |
| MEG_2357\|Drugs\|betalactams\|Class_A_betalactamases\|CTX | CTX | ESBL |
| MEG_2358\|Drugs\|betalactams\|Class_A_betalactamases\|CTX | CTX | ESBL |
| MEG_2359\|Drugs\|betalactams\|Class_A_betalactamases\|CTX | CTX | ESBL |
| MEG_2360\|Drugs\|betalactams\|Class_A_betalactamases\|CTX | CTX | ESBL |
| MEG_2361\|Drugs\|betalactams\|Class_A_betalactamases\|CTX | CTX | ESBL |
| MEG_2362\|Drugs\|betalactams\|Class_A_betalactamases\|CTX | CTX | ESBL |
| MEG_2363\|Drugs\|betalactams\|Class_A_betalactamases\|CTX | CTX | ESBL |
| MEG_2364\|Drugs\|betalactams\|Class_A_betalactamases\|CTX | CTX | ESBL |
| MEG_2365\|Drugs\|betalactams\|Class_A_betalactamases\|CTX | CTX | ESBL |
| MEG_2366\|Drugs\|betalactams\|Class_A_betalactamases\|CTX | CTX | ESBL |
| MEG_2367\|Drugs\|betalactams\|Class_A_betalactamases\|CTX | CTX | ESBL |
| MEG_2368\|Drugs\|betalactams\|Class_A_betalactamases\|CTX | CTX | ESBL |
| MEG_2369\|Drugs\|betalactams\|Class_A_betalactamases\|CTX | CTX | ESBL |
| MEG_2370\|Drugs\|betalactams\|Class_A_betalactamases\|CTX | CTX | ESBL |
| MEG_2371\|Drugs\|betalactams\|Class_A_betalactamases\|CTX | CTX | ESBL |
| MEG_2372\|Drugs\|betalactams\|Class_A_betalactamases\|CTX | CTX | ESBL |
| MEG_2373\|Drugs\|betalactams\|Class_A_betalactamases\|CTX | CTX | ESBL |
| MEG_2374\|Drugs\|betalactams\|Class_A_betalactamases\|CTX | CTX | ESBL |
| MEG_2375\|Drugs\|betalactams\|Class_A_betalactamases\|CTX | CTX | ESBL |
| MEG_2376\|Drugs\|betalactams\|Class_A_betalactamases\|CTX | CTX | ESBL |
| MEG_2377\|Drugs\|betalactams\|Class_A_betalactamases\|CTX | CTX | ESBL |
| MEG_2378\|Drugs\|betalactams\|Class_A_betalactamases\|CTX | CTX | ESBL |
| MEG_2379\|Drugs\|betalactams\|Class_A_betalactamases\|CTX | CTX | ESBL |
| MEG_2380\|Drugs\|betalactams\|Class_A_betalactamases\|CTX | CTX | ESBL |
| MEG_2381\|Drugs\|betalactams\|Class_A_betalactamases\|CTX | CTX | ESBL |
| MEG_2382\|Drugs\|betalactams\|Class_A_betalactamases\|CTX | CTX | ESBL |
| MEG_2383\|Drugs\|betalactams\|Class_A_betalactamases\|CTX | CTX | ESBL |
| MEG_2384\|Drugs\|betalactams\|Class_A_betalactamases\|CTX | CTX | ESBL |
| MEG_2385\|Drugs\|betalactams\|Class_A_betalactamases\|CTX | CTX | ESBL |
| MEG_2386\|Drugs\|betalactams\|Class_A_betalactamases\|CTX | CTX | ESBL |
| MEG_2387\|Drugs\|betalactams\|Class_A_betalactamases\|CTX | CTX | ESBL |
| MEG_2388\|Drugs\|betalactams\|Class_A_betalactamases\|CTX | CTX | ESBL |
| MEG_2389\|Drugs\|betalactams\|Class_A_betalactamases\|CTX | CTX | ESBL |
| MEG_2390\|Drugs\|betalactams\|Class_A_betalactamases\|CTX | CTX | ESBL |
| MEG_2391\|Drugs\|betalactams\|Class_A_betalactamases\|CTX | CTX | ESBL |
| MEG_2392\|Drugs\|betalactams\|Class_A_betalactamases\|CTX | CTX | ESBL |
| MEG_2393\|Drugs\|betalactams\|Class_A_betalactamases\|CTX | CTX | ESBL |
| MEG_2394\|Drugs\|betalactams\|Class_A_betalactamases\|CTX | CTX | ESBL |
| MEG_2395\|Drugs\|betalactams\|Class_A_betalactamases\|CTX | CTX | ESBL |
| MEG_2396\|Drugs\|betalactams\|Class_A_betalactamases\|CTX | CTX | ESBL |
| MEG_2397\|Drugs\|betalactams\|Class_A_betalactamases\|CTX | CTX | ESBL |
| MEG_2398\|Drugs\|betalactams\|Class_A_betalactamases\|CTX | CTX | ESBL |
| MEG_2399\|Drugs\|betalactams\|Class_A_betalactamases\|CTX | CTX | ESBL |
| MEG_2400\|Drugs\|betalactams\|Class_A_betalactamases\|CTX | CTX | ESBL |
| MEG_2401\|Drugs\|betalactams\|Class_A_betalactamases\|CTX | CTX | ESBL |
| MEG_2402\|Drugs\|betalactams\|Class_A_betalactamases\|CTX | CTX | ESBL |
| MEG_2403\|Drugs\|betalactams\|Class_A_betalactamases\|CTX | CTX | ESBL |
| MEG_2404\|Drugs\|betalactams\|Class_A_betalactamases\|CTX | CTX | ESBL |
| MEG_2405\|Drugs\|betalactams\|Class_A_betalactamases\|CTX | CTX | ESBL |
| MEG_2406\|Drugs\|betalactams\|Class_A_betalactamases\|CTX | CTX | ESBL |
| MEG_2407\|Drugs\|betalactams\|Class_A_betalactamases\|CTX | CTX | ESBL |
| MEG_2408\|Drugs\|betalactams\|Class_A_betalactamases\|CTX | CTX | ESBL |
| MEG_2409\|Drugs\|betalactams\|Class_A_betalactamases\|CTX | CTX | ESBL |
| MEG_2410\|Drugs\|betalactams\|Class_A_betalactamases\|CTX | CTX | ESBL |
| MEG_2411\|Drugs\|betalactams\|Class_A_betalactamases\|CTX | CTX | ESBL |
| MEG_2412\|Drugs\|betalactams\|Class_A_betalactamases\|CTX | CTX | ESBL |
| MEG_2413\|Drugs\|betalactams\|Class_A_betalactamases\|CTX | CTX | ESBL |
| MEG_2414\|Drugs\|betalactams\|Class_A_betalactamases\|CTX | CTX | ESBL |
| MEG_2415\|Drugs\|betalactams\|Class_A_betalactamases\|CTX | CTX | ESBL |
| MEG_2416\|Drugs\|betalactams\|Class_A_betalactamases\|CTX | CTX | ESBL |
| MEG_2417\|Drugs\|betalactams\|Class_A_betalactamases\|CTX | CTX | ESBL |
| MEG_2418\|Drugs\|betalactams\|Class_A_betalactamases\|CTX | CTX | ESBL |
| MEG_2419\|Drugs\|betalactams\|Class_A_betalactamases\|CTX | CTX | ESBL |
| MEG_2420\|Drugs\|betalactams\|Class_A_betalactamases\|CTX | CTX | ESBL |
| MEG_2421\|Drugs\|betalactams\|Class_A_betalactamases\|CTX | CTX | ESBL |
| MEG_2422\|Drugs\|betalactams\|Class_A_betalactamases\|CTX | CTX | ESBL |
| MEG_2423\|Drugs\|betalactams\|Class_A_betalactamases\|CTX | CTX | ESBL |
| MEG_2424\|Drugs\|betalactams\|Class_A_betalactamases\|CTX | CTX | ESBL |
| MEG_2425\|Drugs\|betalactams\|Class_A_betalactamases\|CTX | CTX | ESBL |
| MEG_2426\|Drugs\|betalactams\|Class_A_betalactamases\|CTX | CTX | ESBL |
| MEG_2427\|Drugs\|betalactams\|Class_A_betalactamases\|CTX | CTX | ESBL |
| MEG_2428\|Drugs\|betalactams\|Class_A_betalactamases\|CTX | CTX | ESBL |
| MEG_2429\|Drugs\|betalactams\|Class_A_betalactamases\|CTX | CTX | ESBL |
| MEG_2430\|Drugs\|betalactams\|Class_A_betalactamases\|CTX | CTX | ESBL |
| MEG_2431\|Drugs\|betalactams\|Class_A_betalactamases\|CTX | CTX | ESBL |
| MEG_2432\|Drugs\|betalactams\|Class_A_betalactamases\|CTX | CTX | ESBL |
| MEG_2433\|Drugs\|betalactams\|Class_A_betalactamases\|CTX | CTX | ESBL |
| MEG_2434\|Drugs\|betalactams\|Class_A_betalactamases\|CTX | CTX | ESBL |
| MEG_2435\|Drugs\|betalactams\|Class_A_betalactamases\|CTX | CTX | ESBL |
| MEG_2436\|Drugs\|betalactams\|Class_A_betalactamases\|CTX | CTX | ESBL |
| MEG_2437\|Drugs\|betalactams\|Class_A_betalactamases\|CTX | CTX | ESBL |
| MEG_2438\|Drugs\|betalactams\|Class_A_betalactamases\|CTX | CTX | ESBL |
| MEG_8696\|Drugs\|betalactams\|Class_A_betalactamases\|SHV | SHV | ESBL |
| MEG_8697\|Drugs\|betalactams\|Class_A_betalactamases\|SHV | SHV | ESBL |
| MEG_6188\|Drugs\|betalactams\|Class_A_betalactamases\|SHV | SHV | ESBL |
| MEG_6189\|Drugs\|betalactams\|Class_A_betalactamases\|SHV | SHV | ESBL |
| MEG_6190\|Drugs\|betalactams\|Class_A_betalactamases\|SHV | SHV | ESBL |
| MEG_6191\|Drugs\|betalactams\|Class_A_betalactamases\|SHV | SHV | ESBL |
| MEG_6192\|Drugs\|betalactams\|Class_A_betalactamases\|SHV | SHV | ESBL |
| MEG_6193\|Drugs\|betalactams\|Class_A_betalactamases\|SHV | SHV | ESBL |
| MEG_6194\|Drugs\|betalactams\|Class_A_betalactamases\|SHV | SHV | ESBL |
| MEG_6195\|Drugs\|betalactams\|Class_A_betalactamases\|SHV | SHV | ESBL |
| MEG_6199\|Drugs\|betalactams\|Class_A_betalactamases\|SHV | SHV | ESBL |
| MEG_6202\|Drugs\|betalactams\|Class_A_betalactamases\|SHV | SHV | ESBL |
| MEG_6203\|Drugs\|betalactams\|Class_A_betalactamases\|SHV | SHV | ESBL |
| MEG_6205\|Drugs\|betalactams\|Class_A_betalactamases\|SHV | SHV | ESBL |
| MEG_6206\|Drugs\|betalactams\|Class_A_betalactamases\|SHV | SHV | ESBL |
| MEG_6207\|Drugs\|betalactams\|Class_A_betalactamases\|SHV | SHV | ESBL |
| MEG_6208\|Drugs\|betalactams\|Class_A_betalactamases\|SHV | SHV | ESBL |
| MEG_6209\|Drugs\|betalactams\|Class_A_betalactamases\|SHV | SHV | ESBL |
| MEG_6210\|Drugs\|betalactams\|Class_A_betalactamases\|SHV | SHV | ESBL |
| MEG_6211\|Drugs\|betalactams\|Class_A_betalactamases\|SHV | SHV | ESBL |
| MEG_6212\|Drugs\|betalactams\|Class_A_betalactamases\|SHV | SHV | ESBL |
| MEG_6213\|Drugs\|betalactams\|Class_A_betalactamases\|SHV | SHV | ESBL |
| MEG_6215\|Drugs\|betalactams\|Class_A_betalactamases\|SHV | SHV | ESBL |
| MEG_6217\|Drugs\|betalactams\|Class_A_betalactamases\|SHV | SHV | ESBL |
| MEG_6218\|Drugs\|betalactams\|Class_A_betalactamases\|SHV | SHV | ESBL |
| MEG_6219\|Drugs\|betalactams\|Class_A_betalactamases\|SHV | SHV | ESBL |
| MEG_6220\|Drugs\|betalactams\|Class_A_betalactamases\|SHV | SHV | ESBL |
| MEG_6221\|Drugs\|betalactams\|Class_A_betalactamases\|SHV | SHV | ESBL |
| MEG_6222\|Drugs\|betalactams\|Class_A_betalactamases\|SHV | SHV | ESBL |
| MEG_6223\|Drugs\|betalactams\|Class_A_betalactamases\|SHV | SHV | ESBL |
| MEG_6225\|Drugs\|betalactams\|Class_A_betalactamases\|SHV | SHV | ESBL |
| MEG_6226\|Drugs\|betalactams\|Class_A_betalactamases\|SHV | SHV | ESBL |
| MEG_6227\|Drugs\|betalactams\|Class_A_betalactamases\|SHV | SHV | ESBL |
| MEG_6228\|Drugs\|betalactams\|Class_A_betalactamases\|SHV | SHV | ESBL |
| MEG_6229\|Drugs\|betalactams\|Class_A_betalactamases\|SHV | SHV | ESBL |
| MEG_6230\|Drugs\|betalactams\|Class_A_betalactamases\|SHV | SHV | ESBL |
| MEG_6232\|Drugs\|betalactams\|Class_A_betalactamases\|SHV | SHV | ESBL |
| MEG_6234\|Drugs\|betalactams\|Class_A_betalactamases\|SHV | SHV | ESBL |
| MEG_6235\|Drugs\|betalactams\|Class_A_betalactamases\|SHV | SHV | ESBL |
| MEG_6236\|Drugs\|betalactams\|Class_A_betalactamases\|SHV | SHV | ESBL |
| MEG_6237\|Drugs\|betalactams\|Class_A_betalactamases\|SHV | SHV | ESBL |
| MEG_6238\|Drugs\|betalactams\|Class_A_betalactamases\|SHV | SHV | ESBL |
| MEG_6239\|Drugs\|betalactams\|Class_A_betalactamases\|SHV | SHV | ESBL |
| MEG_6243\|Drugs\|betalactams\|Class_A_betalactamases\|SHV | SHV | ESBL |
| MEG_6244\|Drugs\|betalactams\|Class_A_betalactamases\|SHV | SHV | ESBL |
| MEG_6245\|Drugs\|betalactams\|Class_A_betalactamases\|SHV | SHV | ESBL |
| MEG_6246\|Drugs\|betalactams\|Class_A_betalactamases\|SHV | SHV | ESBL |
| MEG_6247\|Drugs\|betalactams\|Class_A_betalactamases\|SHV | SHV | ESBL |
| MEG_6248\|Drugs\|betalactams\|Class_A_betalactamases\|SHV | SHV | ESBL |
| MEG_6249\|Drugs\|betalactams\|Class_A_betalactamases\|SHV | SHV | ESBL |
| MEG_6250\|Drugs\|betalactams\|Class_A_betalactamases\|SHV | SHV | ESBL |
| MEG_6251\|Drugs\|betalactams\|Class_A_betalactamases\|SHV | SHV | ESBL |
| MEG_6252\|Drugs\|betalactams\|Class_A_betalactamases\|SHV | SHV | ESBL |
| MEG_6253\|Drugs\|betalactams\|Class_A_betalactamases\|SHV | SHV | ESBL |
| MEG_6254\|Drugs\|betalactams\|Class_A_betalactamases\|SHV | SHV | ESBL |
| MEG_6255\|Drugs\|betalactams\|Class_A_betalactamases\|SHV | SHV | ESBL |
| MEG_6256\|Drugs\|betalactams\|Class_A_betalactamases\|SHV | SHV | ESBL |
| MEG_6257\|Drugs\|betalactams\|Class_A_betalactamases\|SHV | SHV | ESBL |
| MEG_6258\|Drugs\|betalactams\|Class_A_betalactamases\|SHV | SHV | ESBL |
| MEG_6259\|Drugs\|betalactams\|Class_A_betalactamases\|SHV | SHV | ESBL |
| MEG_6260\|Drugs\|betalactams\|Class_A_betalactamases\|SHV | SHV | ESBL |
| MEG_6261\|Drugs\|betalactams\|Class_A_betalactamases\|SHV | SHV | ESBL |
| MEG_6262\|Drugs\|betalactams\|Class_A_betalactamases\|SHV | SHV | ESBL |
| MEG_6263\|Drugs\|betalactams\|Class_A_betalactamases\|SHV | SHV | ESBL |
| MEG_6264\|Drugs\|betalactams\|Class_A_betalactamases\|SHV | SHV | ESBL |
| MEG_6265\|Drugs\|betalactams\|Class_A_betalactamases\|SHV | SHV | ESBL |
| MEG_6266\|Drugs\|betalactams\|Class_A_betalactamases\|SHV | SHV | ESBL |
| MEG_6267\|Drugs\|betalactams\|Class_A_betalactamases\|SHV | SHV | ESBL |
| MEG_6268\|Drugs\|betalactams\|Class_A_betalactamases\|SHV | SHV | ESBL |
| MEG_6269\|Drugs\|betalactams\|Class_A_betalactamases\|SHV | SHV | ESBL |
| MEG_6270\|Drugs\|betalactams\|Class_A_betalactamases\|SHV | SHV | ESBL |
| MEG_6271\|Drugs\|betalactams\|Class_A_betalactamases\|SHV | SHV | ESBL |
| MEG_6272\|Drugs\|betalactams\|Class_A_betalactamases\|SHV | SHV | ESBL |
| MEG_6273\|Drugs\|betalactams\|Class_A_betalactamases\|SHV | SHV | ESBL |
| MEG_6274\|Drugs\|betalactams\|Class_A_betalactamases\|SHV | SHV | ESBL |
| MEG_6275\|Drugs\|betalactams\|Class_A_betalactamases\|SHV | SHV | ESBL |
| MEG_6276\|Drugs\|betalactams\|Class_A_betalactamases\|SHV | SHV | ESBL |
| MEG_6277\|Drugs\|betalactams\|Class_A_betalactamases\|SHV | SHV | ESBL |
| MEG_6278\|Drugs\|betalactams\|Class_A_betalactamases\|SHV | SHV | ESBL |
| MEG_6279\|Drugs\|betalactams\|Class_A_betalactamases\|SHV | SHV | ESBL |
| MEG_6280\|Drugs\|betalactams\|Class_A_betalactamases\|SHV | SHV | ESBL |
| MEG_6281\|Drugs\|betalactams\|Class_A_betalactamases\|SHV | SHV | ESBL |
| MEG_6282\|Drugs\|betalactams\|Class_A_betalactamases\|SHV | SHV | ESBL |
| MEG_6283\|Drugs\|betalactams\|Class_A_betalactamases\|SHV | SHV | ESBL |
| MEG_6284\|Drugs\|betalactams\|Class_A_betalactamases\|SHV | SHV | ESBL |
| MEG_6285\|Drugs\|betalactams\|Class_A_betalactamases\|SHV | SHV | ESBL |
| MEG_6286\|Drugs\|betalactams\|Class_A_betalactamases\|SHV | SHV | ESBL |
| MEG_6287\|Drugs\|betalactams\|Class_A_betalactamases\|SHV | SHV | ESBL |
| MEG_6288\|Drugs\|betalactams\|Class_A_betalactamases\|SHV | SHV | ESBL |
| MEG_6289\|Drugs\|betalactams\|Class_A_betalactamases\|SHV | SHV | ESBL |
| MEG_6290\|Drugs\|betalactams\|Class_A_betalactamases\|SHV | SHV | ESBL |
| MEG_6291\|Drugs\|betalactams\|Class_A_betalactamases\|SHV | SHV | ESBL |
| MEG_6292\|Drugs\|betalactams\|Class_A_betalactamases\|SHV | SHV | ESBL |
| MEG_6293\|Drugs\|betalactams\|Class_A_betalactamases\|SHV | SHV | ESBL |
| MEG_6294\|Drugs\|betalactams\|Class_A_betalactamases\|SHV | SHV | ESBL |
| MEG_6295\|Drugs\|betalactams\|Class_A_betalactamases\|SHV | SHV | ESBL |
| MEG_6296\|Drugs\|betalactams\|Class_A_betalactamases\|SHV | SHV | ESBL |
| MEG_6297\|Drugs\|betalactams\|Class_A_betalactamases\|SHV | SHV | ESBL |
| MEG_6298\|Drugs\|betalactams\|Class_A_betalactamases\|SHV | SHV | ESBL |
| MEG_6299\|Drugs\|betalactams\|Class_A_betalactamases\|SHV | SHV | ESBL |
| MEG_6300\|Drugs\|betalactams\|Class_A_betalactamases\|SHV | SHV | ESBL |
| MEG_6301\|Drugs\|betalactams\|Class_A_betalactamases\|SHV | SHV | ESBL |
| MEG_6302\|Drugs\|betalactams\|Class_A_betalactamases\|SHV | SHV | ESBL |
| MEG_6303\|Drugs\|betalactams\|Class_A_betalactamases\|SHV | SHV | ESBL |
| MEG_6304\|Drugs\|betalactams\|Class_A_betalactamases\|SHV | SHV | ESBL |
| MEG_6305\|Drugs\|betalactams\|Class_A_betalactamases\|SHV | SHV | ESBL |
| MEG_6306\|Drugs\|betalactams\|Class_A_betalactamases\|SHV | SHV | ESBL |
| MEG_6307\|Drugs\|betalactams\|Class_A_betalactamases\|SHV | SHV | ESBL |
| MEG_6308\|Drugs\|betalactams\|Class_A_betalactamases\|SHV | SHV | ESBL |
| MEG_6309\|Drugs\|betalactams\|Class_A_betalactamases\|SHV | SHV | ESBL |
| MEG_6310\|Drugs\|betalactams\|Class_A_betalactamases\|SHV | SHV | ESBL |
| MEG_6311\|Drugs\|betalactams\|Class_A_betalactamases\|SHV | SHV | ESBL |
| MEG_6312\|Drugs\|betalactams\|Class_A_betalactamases\|SHV | SHV | ESBL |
| MEG_6313\|Drugs\|betalactams\|Class_A_betalactamases\|SHV | SHV | ESBL |
| MEG_6314\|Drugs\|betalactams\|Class_A_betalactamases\|SHV | SHV | ESBL |
| MEG_6315\|Drugs\|betalactams\|Class_A_betalactamases\|SHV | SHV | ESBL |
| MEG_6316\|Drugs\|betalactams\|Class_A_betalactamases\|SHV | SHV | ESBL |
| MEG_6317\|Drugs\|betalactams\|Class_A_betalactamases\|SHV | SHV | ESBL |
| MEG_6318\|Drugs\|betalactams\|Class_A_betalactamases\|SHV | SHV | ESBL |
| MEG_6319\|Drugs\|betalactams\|Class_A_betalactamases\|SHV | SHV | ESBL |
| MEG_6320\|Drugs\|betalactams\|Class_A_betalactamases\|SHV | SHV | ESBL |
| MEG_6321\|Drugs\|betalactams\|Class_A_betalactamases\|SHV | SHV | ESBL |
| MEG_6322\|Drugs\|betalactams\|Class_A_betalactamases\|SHV | SHV | ESBL |
| MEG_6323\|Drugs\|betalactams\|Class_A_betalactamases\|SHV | SHV | ESBL |
| MEG_6324\|Drugs\|betalactams\|Class_A_betalactamases\|SHV | SHV | ESBL |
| MEG_6325\|Drugs\|betalactams\|Class_A_betalactamases\|SHV | SHV | ESBL |
| MEG_6326\|Drugs\|betalactams\|Class_A_betalactamases\|SHV | SHV | ESBL |
| MEG_6327\|Drugs\|betalactams\|Class_A_betalactamases\|SHV | SHV | ESBL |
| MEG_6328\|Drugs\|betalactams\|Class_A_betalactamases\|SHV | SHV | ESBL |
| MEG_6329\|Drugs\|betalactams\|Class_A_betalactamases\|SHV | SHV | ESBL |
| MEG_6330\|Drugs\|betalactams\|Class_A_betalactamases\|SHV | SHV | ESBL |
| MEG_6331\|Drugs\|betalactams\|Class_A_betalactamases\|SHV | SHV | ESBL |
| MEG_6332\|Drugs\|betalactams\|Class_A_betalactamases\|SHV | SHV | ESBL |
| MEG_6333\|Drugs\|betalactams\|Class_A_betalactamases\|SHV | SHV | ESBL |
| MEG_6334\|Drugs\|betalactams\|Class_A_betalactamases\|SHV | SHV | ESBL |
| MEG_6335\|Drugs\|betalactams\|Class_A_betalactamases\|SHV | SHV | ESBL |
| MEG_6336\|Drugs\|betalactams\|Class_A_betalactamases\|SHV | SHV | ESBL |
| MEG_6337\|Drugs\|betalactams\|Class_A_betalactamases\|SHV | SHV | ESBL |
| MEG_6338\|Drugs\|betalactams\|Class_A_betalactamases\|SHV | SHV | ESBL |
| MEG_6339\|Drugs\|betalactams\|Class_A_betalactamases\|SHV | SHV | ESBL |
| MEG_6340\|Drugs\|betalactams\|Class_A_betalactamases\|SHV | SHV | ESBL |
| MEG_6341\|Drugs\|betalactams\|Class_A_betalactamases\|SHV | SHV | ESBL |
| MEG_6342\|Drugs\|betalactams\|Class_A_betalactamases\|SHV | SHV | ESBL |
| MEG_6343\|Drugs\|betalactams\|Class_A_betalactamases\|SHV | SHV | ESBL |
| MEG_6344\|Drugs\|betalactams\|Class_A_betalactamases\|SHV | SHV | ESBL |
| MEG_6345\|Drugs\|betalactams\|Class_A_betalactamases\|SHV | SHV | ESBL |
| MEG_6346\|Drugs\|betalactams\|Class_A_betalactamases\|SHV | SHV | ESBL |
| MEG_6347\|Drugs\|betalactams\|Class_A_betalactamases\|SHV | SHV | ESBL |
| MEG_6348\|Drugs\|betalactams\|Class_A_betalactamases\|SHV | SHV | ESBL |
| MEG_6349\|Drugs\|betalactams\|Class_A_betalactamases\|SHV | SHV | ESBL |
| MEG_6350\|Drugs\|betalactams\|Class_A_betalactamases\|SHV | SHV | ESBL |
| MEG_6351\|Drugs\|betalactams\|Class_A_betalactamases\|SHV | SHV | ESBL |
| MEG_6352\|Drugs\|betalactams\|Class_A_betalactamases\|SHV | SHV | ESBL |
| MEG_6353\|Drugs\|betalactams\|Class_A_betalactamases\|SHV | SHV | ESBL |
| MEG_6354\|Drugs\|betalactams\|Class_A_betalactamases\|SHV | SHV | ESBL |
| MEG_6355\|Drugs\|betalactams\|Class_A_betalactamases\|SHV | SHV | ESBL |
| MEG_6356\|Drugs\|betalactams\|Class_A_betalactamases\|SHV | SHV | ESBL |
| MEG_6357\|Drugs\|betalactams\|Class_A_betalactamases\|SHV | SHV | ESBL |
| MEG_6358\|Drugs\|betalactams\|Class_A_betalactamases\|SHV | SHV | ESBL |
| MEG_6359\|Drugs\|betalactams\|Class_A_betalactamases\|SHV | SHV | ESBL |
| MEG_6360\|Drugs\|betalactams\|Class_A_betalactamases\|SHV | SHV | ESBL |
| MEG_6361\|Drugs\|betalactams\|Class_A_betalactamases\|SHV | SHV | ESBL |
| MEG_6362\|Drugs\|betalactams\|Class_A_betalactamases\|SHV | SHV | ESBL |
| MEG_6363\|Drugs\|betalactams\|Class_A_betalactamases\|SHV | SHV | ESBL |
| MEG_6364\|Drugs\|betalactams\|Class_A_betalactamases\|SHV | SHV | ESBL |
| MEG_6365\|Drugs\|betalactams\|Class_A_betalactamases\|SHV | SHV | ESBL |
| MEG_6366\|Drugs\|betalactams\|Class_A_betalactamases\|SHV | SHV | ESBL |
| MEG_6367\|Drugs\|betalactams\|Class_A_betalactamases\|SHV | SHV | ESBL |
| MEG_6368\|Drugs\|betalactams\|Class_A_betalactamases\|SHV | SHV | ESBL |
| MEG_6369\|Drugs\|betalactams\|Class_A_betalactamases\|SHV | SHV | ESBL |
| MEG_6370\|Drugs\|betalactams\|Class_A_betalactamases\|SHV | SHV | ESBL |
| MEG_6371\|Drugs\|betalactams\|Class_A_betalactamases\|SHV | SHV | ESBL |
| MEG_6372\|Drugs\|betalactams\|Class_A_betalactamases\|SHV | SHV | ESBL |
| MEG_6373\|Drugs\|betalactams\|Class_A_betalactamases\|SHV | SHV | ESBL |
| MEG_6374\|Drugs\|betalactams\|Class_A_betalactamases\|SHV | SHV | ESBL |
| MEG_6375\|Drugs\|betalactams\|Class_A_betalactamases\|SHV | SHV | ESBL |
| MEG_6376\|Drugs\|betalactams\|Class_A_betalactamases\|SHV | SHV | ESBL |
| MEG_6377\|Drugs\|betalactams\|Class_A_betalactamases\|SHV | SHV | ESBL |
| MEG_6378\|Drugs\|betalactams\|Class_A_betalactamases\|SHV | SHV | ESBL |
| MEG_6379\|Drugs\|betalactams\|Class_A_betalactamases\|SHV | SHV | ESBL |
| MEG_6380\|Drugs\|betalactams\|Class_A_betalactamases\|SHV | SHV | ESBL |
| MEG_6381\|Drugs\|betalactams\|Class_A_betalactamases\|SHV | SHV | ESBL |
| MEG_6382\|Drugs\|betalactams\|Class_A_betalactamases\|SHV | SHV | ESBL |
| MEG_6383\|Drugs\|betalactams\|Class_A_betalactamases\|SHV | SHV | ESBL |
| MEG_6384\|Drugs\|betalactams\|Class_A_betalactamases\|SHV | SHV | ESBL |
| MEG_6385\|Drugs\|betalactams\|Class_A_betalactamases\|SHV | SHV | ESBL |
| MEG_6386\|Drugs\|betalactams\|Class_A_betalactamases\|SHV | SHV | ESBL |
| MEG_6387\|Drugs\|betalactams\|Class_A_betalactamases\|SHV | SHV | ESBL |
| MEG_6388\|Drugs\|betalactams\|Class_A_betalactamases\|SHV | SHV | ESBL |
| MEG_6389\|Drugs\|betalactams\|Class_A_betalactamases\|SHV | SHV | ESBL |
| MEG_6390\|Drugs\|betalactams\|Class_A_betalactamases\|SHV | SHV | ESBL |
| MEG_6391\|Drugs\|betalactams\|Class_A_betalactamases\|SHV | SHV | ESBL |
| MEG_6392\|Drugs\|betalactams\|Class_A_betalactamases\|SHV | SHV | ESBL |
| MEG_6393\|Drugs\|betalactams\|Class_A_betalactamases\|SHV | SHV | ESBL |
| MEG_6394\|Drugs\|betalactams\|Class_A_betalactamases\|SHV | SHV | ESBL |
| MEG_6395\|Drugs\|betalactams\|Class_A_betalactamases\|SHV | SHV | ESBL |
| MEG_6396\|Drugs\|betalactams\|Class_A_betalactamases\|SHV | SHV | ESBL |
| MEG_6397\|Drugs\|betalactams\|Class_A_betalactamases\|SHV | SHV | ESBL |
| MEG_6398\|Drugs\|betalactams\|Class_A_betalactamases\|SHV | SHV | ESBL |
| MEG_6399\|Drugs\|betalactams\|Class_A_betalactamases\|SHV | SHV | ESBL |
| MEG_6400\|Drugs\|betalactams\|Class_A_betalactamases\|SHV | SHV | ESBL |
| MEG_6401\|Drugs\|betalactams\|Class_A_betalactamases\|SHV | SHV | ESBL |
| MEG_6402\|Drugs\|betalactams\|Class_A_betalactamases\|SHV | SHV | ESBL |
| MEG_6404\|Drugs\|betalactams\|Class_A_betalactamases\|SHV | SHV | ESBL |
| MEG_6405\|Drugs\|betalactams\|Class_A_betalactamases\|SHV | SHV | ESBL |
| MEG_6406\|Drugs\|betalactams\|Class_A_betalactamases\|SHV | SHV | ESBL |
| MEG_6407\|Drugs\|betalactams\|Class_A_betalactamases\|SHV | SHV | ESBL |
| MEG_6408\|Drugs\|betalactams\|Class_A_betalactamases\|SHV | SHV | ESBL |
| MEG_6409\|Drugs\|betalactams\|Class_A_betalactamases\|SHV | SHV | ESBL |
| MEG_6410\|Drugs\|betalactams\|Class_A_betalactamases\|SHV | SHV | ESBL |
| MEG_6411\|Drugs\|betalactams\|Class_A_betalactamases\|SHV | SHV | ESBL |
| MEG_6412\|Drugs\|betalactams\|Class_A_betalactamases\|SHV | SHV | ESBL |
| MEG_6413\|Drugs\|betalactams\|Class_A_betalactamases\|SHV | SHV | ESBL |
| MEG_6414\|Drugs\|betalactams\|Class_A_betalactamases\|SHV | SHV | ESBL |
| MEG_6415\|Drugs\|betalactams\|Class_A_betalactamases\|SHV | SHV | ESBL |
| MEG_6416\|Drugs\|betalactams\|Class_A_betalactamases\|SHV | SHV | ESBL |
| MEG_6417\|Drugs\|betalactams\|Class_A_betalactamases\|SHV | SHV | ESBL |
| MEG_6418\|Drugs\|betalactams\|Class_A_betalactamases\|SHV | SHV | ESBL |
| MEG_6419\|Drugs\|betalactams\|Class_A_betalactamases\|SHV | SHV | ESBL |
| MEG_6420\|Drugs\|betalactams\|Class_A_betalactamases\|SHV | SHV | ESBL |
| MEG_6421\|Drugs\|betalactams\|Class_A_betalactamases\|SHV | SHV | ESBL |
| MEG_6422\|Drugs\|betalactams\|Class_A_betalactamases\|SHV | SHV | ESBL |
| MEG_6423\|Drugs\|betalactams\|Class_A_betalactamases\|SHV | SHV | ESBL |
| MEG_6424\|Drugs\|betalactams\|Class_A_betalactamases\|SHV | SHV | ESBL |
| MEG_6425\|Drugs\|betalactams\|Class_A_betalactamases\|SHV | SHV | ESBL |
| MEG_6426\|Drugs\|betalactams\|Class_A_betalactamases\|SHV | SHV | ESBL |
| MEG_6427\|Drugs\|betalactams\|Class_A_betalactamases\|SHV | SHV | ESBL |
| MEG_6428\|Drugs\|betalactams\|Class_A_betalactamases\|SHV | SHV | ESBL |
| MEG_6429\|Drugs\|betalactams\|Class_A_betalactamases\|SHV | SHV | ESBL |
| MEG_6430\|Drugs\|betalactams\|Class_A_betalactamases\|SHV | SHV | ESBL |
| MEG_6431\|Drugs\|betalactams\|Class_A_betalactamases\|SHV | SHV | ESBL |
| MEG_6432\|Drugs\|betalactams\|Class_A_betalactamases\|SHV | SHV | ESBL |
| MEG_6433\|Drugs\|betalactams\|Class_A_betalactamases\|SHV | SHV | ESBL |
| MEG_6434\|Drugs\|betalactams\|Class_A_betalactamases\|SHV | SHV | ESBL |
| MEG_6435\|Drugs\|betalactams\|Class_A_betalactamases\|SHV | SHV | ESBL |
| MEG_6436\|Drugs\|betalactams\|Class_A_betalactamases\|SHV | SHV | ESBL |
| MEG_6437\|Drugs\|betalactams\|Class_A_betalactamases\|SHV | SHV | ESBL |
| MEG_6438\|Drugs\|betalactams\|Class_A_betalactamases\|SHV | SHV | ESBL |
| MEG_6439\|Drugs\|betalactams\|Class_A_betalactamases\|SHV | SHV | ESBL |
| MEG_6440\|Drugs\|betalactams\|Class_A_betalactamases\|SHV | SHV | ESBL |
| MEG_6441\|Drugs\|betalactams\|Class_A_betalactamases\|SHV | SHV | ESBL |
| MEG_6442\|Drugs\|betalactams\|Class_A_betalactamases\|SHV | SHV | ESBL |
| MEG_6443\|Drugs\|betalactams\|Class_A_betalactamases\|SHV | SHV | ESBL |
| MEG_6444\|Drugs\|betalactams\|Class_A_betalactamases\|SHV | SHV | ESBL |
| MEG_6445\|Drugs\|betalactams\|Class_A_betalactamases\|SHV | SHV | ESBL |
| MEG_6446\|Drugs\|betalactams\|Class_A_betalactamases\|SHV | SHV | ESBL |
| MEG_6447\|Drugs\|betalactams\|Class_A_betalactamases\|SHV | SHV | ESBL |
| MEG_6448\|Drugs\|betalactams\|Class_A_betalactamases\|SHV | SHV | ESBL |
| MEG_6449\|Drugs\|betalactams\|Class_A_betalactamases\|SHV | SHV | ESBL |
| MEG_6450\|Drugs\|betalactams\|Class_A_betalactamases\|SHV | SHV | ESBL |
| MEG_6451\|Drugs\|betalactams\|Class_A_betalactamases\|SHV | SHV | ESBL |
| MEG_6452\|Drugs\|betalactams\|Class_A_betalactamases\|SHV | SHV | ESBL |
| MEG_6453\|Drugs\|betalactams\|Class_A_betalactamases\|SHV | SHV | ESBL |
| MEG_6454\|Drugs\|betalactams\|Class_A_betalactamases\|SHV | SHV | ESBL |
| MEG_6455\|Drugs\|betalactams\|Class_A_betalactamases\|SHV | SHV | ESBL |
| MEG_6456\|Drugs\|betalactams\|Class_A_betalactamases\|SHV | SHV | ESBL |
| MEG_6457\|Drugs\|betalactams\|Class_A_betalactamases\|SHV | SHV | ESBL |
| MEG_6458\|Drugs\|betalactams\|Class_A_betalactamases\|SHV | SHV | ESBL |
| MEG_6459\|Drugs\|betalactams\|Class_A_betalactamases\|SHV | SHV | ESBL |
| MEG_6461\|Drugs\|betalactams\|Class_A_betalactamases\|SHV | SHV | ESBL |
| MEG_6462\|Drugs\|betalactams\|Class_A_betalactamases\|SHV | SHV | ESBL |
| MEG_6463\|Drugs\|betalactams\|Class_A_betalactamases\|SHV | SHV | ESBL |
| MEG_6464\|Drugs\|betalactams\|Class_A_betalactamases\|SHV | SHV | ESBL |
| MEG_6465\|Drugs\|betalactams\|Class_A_betalactamases\|SHV | SHV | ESBL |
| MEG_6466\|Drugs\|betalactams\|Class_A_betalactamases\|SHV | SHV | ESBL |
| MEG_6467\|Drugs\|betalactams\|Class_A_betalactamases\|SHV | SHV | ESBL |
| MEG_6468\|Drugs\|betalactams\|Class_A_betalactamases\|SHV | SHV | ESBL |
| MEG_6469\|Drugs\|betalactams\|Class_A_betalactamases\|SHV | SHV | ESBL |
| MEG_6470\|Drugs\|betalactams\|Class_A_betalactamases\|SHV | SHV | ESBL |
| MEG_6471\|Drugs\|betalactams\|Class_A_betalactamases\|SHV | SHV | ESBL |
| MEG_6472\|Drugs\|betalactams\|Class_A_betalactamases\|SHV | SHV | ESBL |
| MEG_6473\|Drugs\|betalactams\|Class_A_betalactamases\|SHV | SHV | ESBL |
| MEG_6474\|Drugs\|betalactams\|Class_A_betalactamases\|SHV | SHV | ESBL |
| MEG_6475\|Drugs\|betalactams\|Class_A_betalactamases\|SHV | SHV | ESBL |
| MEG_6476\|Drugs\|betalactams\|Class_A_betalactamases\|SHV | SHV | ESBL |
| MEG_6477\|Drugs\|betalactams\|Class_A_betalactamases\|SHV | SHV | ESBL |
| MEG_6478\|Drugs\|betalactams\|Class_A_betalactamases\|SHV | SHV | ESBL |
| MEG_6479\|Drugs\|betalactams\|Class_A_betalactamases\|SHV | SHV | ESBL |
| MEG_6480\|Drugs\|betalactams\|Class_A_betalactamases\|SHV | SHV | ESBL |
| MEG_6481\|Drugs\|betalactams\|Class_A_betalactamases\|SHV | SHV | ESBL |
| MEG_6482\|Drugs\|betalactams\|Class_A_betalactamases\|SHV | SHV | ESBL |
| MEG_6483\|Drugs\|betalactams\|Class_A_betalactamases\|SHV | SHV | ESBL |
| MEG_6484\|Drugs\|betalactams\|Class_A_betalactamases\|SHV | SHV | ESBL |
| MEG_6485\|Drugs\|betalactams\|Class_A_betalactamases\|SHV | SHV | ESBL |
| MEG_6486\|Drugs\|betalactams\|Class_A_betalactamases\|SHV | SHV | ESBL |
| MEG_6487\|Drugs\|betalactams\|Class_A_betalactamases\|SHV | SHV | ESBL |
| MEG_6488\|Drugs\|betalactams\|Class_A_betalactamases\|SHV | SHV | ESBL |
| MEG_6489\|Drugs\|betalactams\|Class_A_betalactamases\|SHV | SHV | ESBL |
| MEG_6490\|Drugs\|betalactams\|Class_A_betalactamases\|SHV | SHV | ESBL |
| MEG_6491\|Drugs\|betalactams\|Class_A_betalactamases\|SHV | SHV | ESBL |
| MEG_6492\|Drugs\|betalactams\|Class_A_betalactamases\|SHV | SHV | ESBL |
| MEG_6493\|Drugs\|betalactams\|Class_A_betalactamases\|SHV | SHV | ESBL |
| MEG_6494\|Drugs\|betalactams\|Class_A_betalactamases\|SHV | SHV | ESBL |
| MEG_6495\|Drugs\|betalactams\|Class_A_betalactamases\|SHV | SHV | ESBL |
| MEG_6496\|Drugs\|betalactams\|Class_A_betalactamases\|SHV | SHV | ESBL |
| MEG_6497\|Drugs\|betalactams\|Class_A_betalactamases\|SHV | SHV | ESBL |
| MEG_6498\|Drugs\|betalactams\|Class_A_betalactamases\|SHV | SHV | ESBL |
